# PORTRAIT: a calibrated patient ‘Passport’ with built-in refusal - describing individuals against a reference population

**DOI:** 10.64898/2026.07.13.26357968

**Authors:** Daniela Oehring

## Abstract

**Background:** Average-based summaries serve individual patients poorly. PORTRAIT is a calibrated, abstention-aware tool that describes where one patient sits relative to a reference population across 12 cardiometabolic markers, how confident that placement is, and which features drive it. PORTRAIT describes; it does not diagnose or predict. Abstention is a designed feature given the known limits of conditional coverage.

**Methods:** Conformal calibration was combined with distribution-free coverage bounds, quantile-regression coordinates, and copula-based joint structure. A frozen reference cohort (n=9421) supplied fixed calibration; a held-out cohort (n=2247) tested transportability across six strata. A release gate required the minimum per-slice coverage to hold across 4 of 5 seeds. Coverage was re-tested under survey weighting to the US adult population. Coherence was reported as a descriptive joint coordinate. Discrimination was summarised with Harrell’s C and multiplicity controlled by BH-FDR. Interface conformance was assessed against defined requirements, Nielsen heuristics, and WCAG 2.2 AA, with attention to automation bias and risk-graph design.

**Results:** Estimating the conformal quantile once on the deep frozen reference held all six strata within band (0.864-0.903) at abstention 0.113, whereas a local re-split - calibrating each stratum on far fewer rows - under-covered to 0.71 at abstention 0.227; the difference is a calibration-depth effect, and coverage survived survey weighting. The release gate passed on 4 of 5 seeds at abstention 0.101 against a nominal 0.90, and in the frozen-reference configuration that ships, all six strata held inside the calibrated band (0.864-0.903). Coherence showed orthogonality 0.444 to raw extremity and correlated 0.892 with a copula-Mahalanobis distance while remaining deliberately non-identical, so it adds per-feature information. Two transfer tests returned negatives; the ocular transfer did not hold coverage at thin-n. Adding coherence changed mortality discrimination by deltaC 0.0047. Interface requirements moved from 14/27/18 to 38/14/7 (MET/PARTIAL/UNMET); Nielsen severity resolved 7 of 10 issues; WCAG 2.2 AA text criteria passed.

**Conclusions:** PORTRAIT situates a patient against a frozen reference, holds coverage under survey weighting to the US adult population, and abstains when calibration cannot be supported. The headline result is a calibration-depth effect: estimating the conformal quantile once on the deep frozen reference held per-stratum coverage on a disjoint cohort.

**Lay summary:** Blood tests are usually read one line at a time, each result checked against a normal range. Two things slip through this net. First, every result can sit inside its own normal range while the combination is genuinely unusual - a pattern no single line flags, so it is never investigated. For the patient that can mean real symptoms left unexplained and a problem recognised later than it should have been: dangerous, not merely inconvenient. Second, when a combined risk score does raise a concern, it rarely shows which results caused it, so a clinician cannot see what to act on. Neither failure is visible on the screen that produced it - nothing tells the reader that something has been missed.

This matters more now that patients can see their own results and, increasingly, paste them into general-purpose artificial-intelligence tools for an interpretation. Those tools return a confident answer whether or not it is justified, and the more markers a person has, the harder the whole picture is to make sense of.

PORTRAIT is built to do the opposite. For one person it describes where each result sits against a large, fixed reference population and how sure that placement is, reporting a range rather than a bare number. It flags which results are surprising given the rest, so the reasons behind a concern are never lost. It gives a single read of how unusual the whole profile is when the results are taken together. And, unlike the displays a clinician sees today, it knows when to stay silent: where a lab report or growth chart shows one line at a time and a risk score gives one number with no breakdown, none of them can tell that the reference population cannot support a reliable statement about a particular person. PORTRAIT refuses in exactly those cases and says so plainly, rather than producing a confident-looking result that is not backed by evidence.

The result is designed to be read on a single page and used in a hospital, a general practice, or a high-street setting, with a plain-language mode for the people whose results are being described. Every statement it makes is checked for calibration against a fixed reference population, and it refuses when that check cannot be met. Looking forward, the same signals that mark a profile as surprising when its results are read together are being examined against real health outcomes, to test whether these patients differ in ways that current displays cannot see.

## 1 Introduction

Clinical results are interpreted against population averages and reference ranges, and this serves the individual patient poorly, because there is no average patient [1]. Approaches that place a person against a modelled reference population have advanced in neuroimaging and lifespan charting [2,3,4], yet routine practice still reduces a multi-marker profile either to per-analyte reference ranges or to a single composite risk score such as QRISK3 [5]. Each discards structure the clinician needs, in opposite ways. A profile that is unusual only in combination - every analyte within its own reference interval, yet the joint pattern rare - produces no out-of-range flag, so it can pass without investigation; for the patient this is a missed or delayed line of enquiry, with real symptoms left unexplained and recognition arriving later than it should. Conversely, a composite score can flag a person without revealing which results drove it [5], leaving the clinician unable to act on it with confidence. Neither failure is visible on the display that produced it, which makes it consequential rather than merely inconvenient.

This gap is widening as patients gain direct access to their own laboratory results [6]. A person can now paste a panel of results into a general-purpose language model and receive a fluent interpretation - one that answers regardless of whether the answer is calibrated or warranted, the automation-bias failure mode long documented for decision-support tools that always respond [7]. The difficulty is compounded because quantitative results are hard to comprehend: numeracy is limited and the presentation format materially changes what is understood [8,9,10]. And because exact conditional coverage cannot be guaranteed in any finite sample [11,12,13], a tool that always answers will, where the reference is thin, answer without warrant. The opportunity, addressed properly, is a description calibrated against a validated reference population that refuses when that backing is absent: it states an individual’s position with demonstrated coverage, or states plainly that the reference is insufficient. That is the gap PORTRAIT fills.

A patient arrives measured on many markers at once. Across 12 cardiometabolic analytes, the clinical question is not what the average value does, but where this individual sits relative to the appropriate reference population, how confident that placement is, which features drive it, and when the placement should not be attempted at all. PORTRAIT answers exactly these four questions. That descriptive scope shapes every design choice that follows.

The motivating difficulty is that there is no average patient [1]. A single label can be shared by hundreds of distinct profiles, so any tool that reduces a heterogeneous individual to one number discards the structure a clinician reads. Two failure modes make this concrete, and they are not the same problem. The first is the jointly-unusual-yet-each-marginal-normal individual: every measurement sits inside its own reference interval, yet the combination is rare. A tool that reports one centile per analyte is silent here, because no single margin is extreme. The second is the opposite: a composite flag fires, but the reader cannot see which features produced it. A single-score tool is silent here, because attribution was discarded on the way to the summary. Per-feature displays preserve attribution but miss joint rarity; single-score displays register joint burden but hide attribution. Neither, alone, is enough.

PORTRAIT holds both on a single clinician-readable page, using three descriptive coordinates plus a principled refusal. The first coordinate is a calibrated per-feature position: where each measurement falls against a conditional reference, reported with a calibrated interval rather than a bare point estimate. The second is per-feature surprise: how extreme each position is, so burden is read directly and attribution is never lost. The third is coherence, a joint coordinate describing how unusual the profile is as a whole. Coherence correlates 0.892 with a copula-Mahalanobis distance but is deliberately not identical to it, and it shows orthogonality 0.444 to raw extremity; that non-redundancy is precisely why it carries per-feature information the margins do not already contain. its a fourth element that governs whether description proceeds at all: a principled refusal, implemented as a visible DESCRIBE / ABSTAIN / REFUSE gate that declines to describe an individual when the frozen reference cannot support a calibrated statement about them. This abstention is treated as a safety feature rather than a limitation. Because exact conditional coverage cannot be guaranteed in any finite sample [11], a tool that always answers will, in the regions where the reference is thin, answer without warrant. The gate makes that failure mode visible and refuses in those regions, which is precisely what makes the tool usable in practice: an individual either receives a description backed by demonstrated coverage, or receives an explicit statement that the reference is insufficient here.

The headline empirical result is stated plainly. When the reference is fitted once on a deep cohort and a disjoint cohort is then scored with no refitting, calibrated coverage holds; when the same data are instead handled by a local re-split, coverage is not held. The single frozen fit is therefore not a convenience but the condition under which the stated coverage is real. This behaviour survives reweighting: after survey weights are applied to represent the US adult population, coverage is retained rather than degraded, so the calibration is a property of the population description and not an artefact of the raw sample composition.

One coordinate warrants a precise description. The coherence coordinate is not the squared Mahalanobis distance and is not a chi-squared statistic on d degrees of freedom. It correlates empirically at about 0.88 with a copula-Mahalanobis distance but is deliberately not identical to it; its orthogonality of about 0.44 to raw extremity is what allows it to carry per-feature information that raw distance does not.

The horizon follows from these two properties together. Because PORTRAIT is calibrated and refuses when it cannot describe, it is packaged as a single offline application with one accessible interface, and its intended descriptive role is the same wherever a complete panel is available - hospital, general practice, or a high-street setting.

## 2 Methods

### 2.1 Overview: from clinical question to a descriptive application

##### Intended use

Intended descriptive role: PORTRAIT positions one individual’s twelve-feature cardiometabolic profile against a fixed reference population (NHANES US adults), reporting calibrated per-feature position, per-feature surprise-given-the-rest, and a joint coherence coordinate, and abstaining or refusing where a pre-specified coverage check on that reference is not met.

Intended input: the complete twelve-feature panel (Table 1) with age and sex. Intended output: a statistical description of position and profile rarity, or an explicit refusal - not a clinical decision.

**Table 1.**
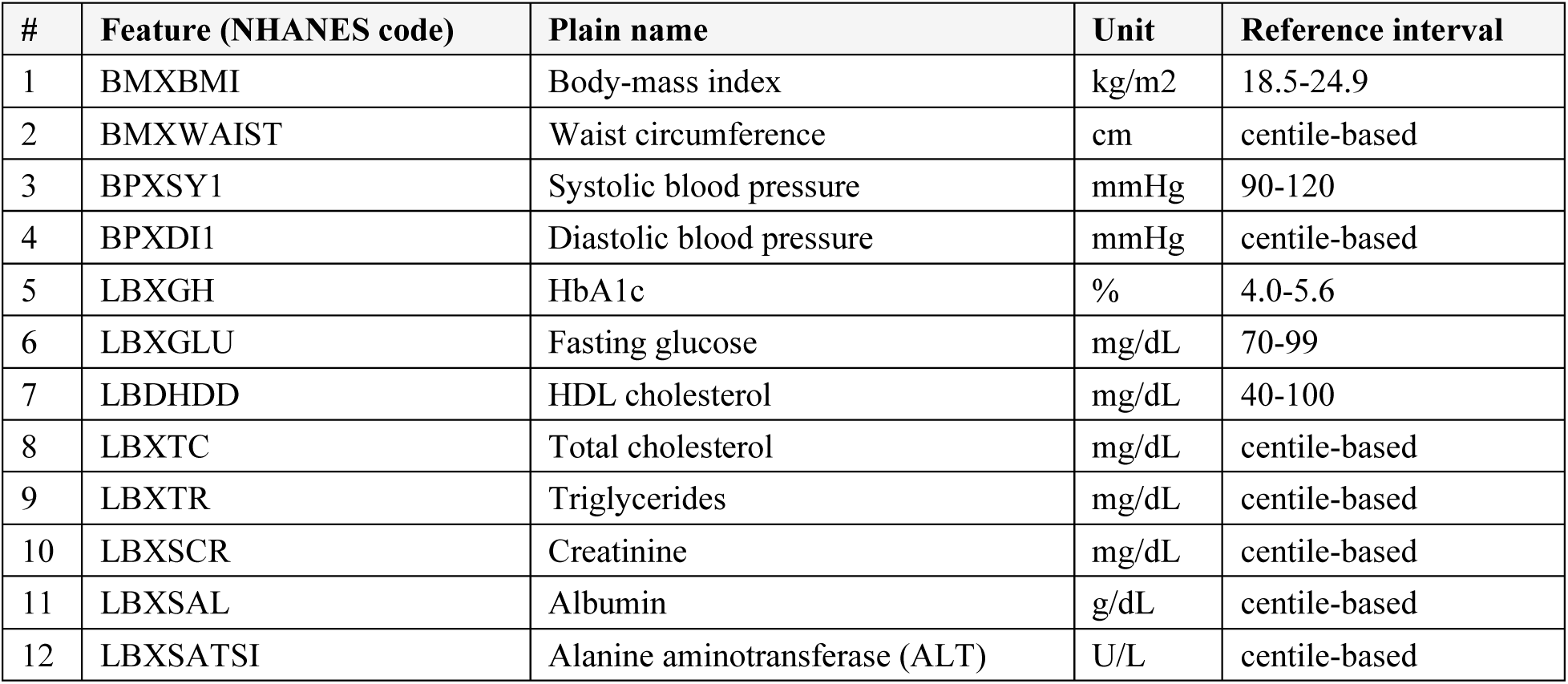
The frozen 12-feature panel. Cardiometabolic and lipid measurements plus age and sex, fixed once as the reference panel. Reference intervals shown where a standard clinical range applies; the remaining features are described by their population centile against the frozen reference rather than a fixed cut-off.

Development proceeded as a staged pipeline. The aim was to produce a calibrated individual description: given one person’s multivariate measurements, a statement of where that individual sits relative to an appropriate reference, how confident that statement is, and which features drive it. The pipeline ran in order: three targeted reviews (mathematical foundations; interface requirements and the regulatory landscape; displays in current clinical use), then derivation of the requirement set and the mathematical spine, then construction of the method, the interface, and a reproducible release. Each review was scoped against a defined question so that its output could be traced forward into a requirement or a design coordinate. The Results are reported in the same order, with substantive findings and tables presented there.

### 2.2 Review of mathematical foundations

The search for existing mathematical solutions to calibrated individual description was scoped as an examination of candidate bodies of theory, each interrogated against a single question: does it, alone, deliver a calibrated, per-feature-attributable, refusal-aware individual description. The theory examined comprised split-conformal prediction [12,13]; distribution-free coverage bounds via the Dvoretzky-Kiefer-Wolfowitz inequality (DKW) [14]; the impossibility of exact conditional coverage [11]; quantile regression [15]; Gaussian copulas [16]; the squared-Mahalanobis / chi-squared-d distance as the classical joint-atypicality scalar; and normative modelling [2,3,4]. Each was assessed for the coordinates it could and could not supply toward calibrated position, per-feature surprise, joint coherence, and a describability gate, and for the preconditions it assumed. The verdict for each is reported in Results 3.1.

### 2.3 Review of interface requirements and the regulatory landscape

Human-factors evidence was gathered across three UX topics (communicating uncertainty and population position; abstention, reject-option and automation bias; clinical dashboard and results display) and graded strong, moderate, or weak, with grades reported as found and no weak finding presented as strong. The regulatory landscape was extracted against named standards: DTAC [17], DCB0129 [18], DCB0160 [19], IEC 62366-1 [20], WCAG 2.2 [21], and the FDA human-factors participant guidance, each mapped to its governing clause and provenance. PORTRAIT is a describe/abstain tool, not a diagnostic device, but these standards were adopted as a shipping bar because hospital procurement and high-street settings require the DTAC/DCB assurance gates and accessible interfaces regardless of device status. The derived requirements are reported in Results 3.2.

### 2.4 Review of displays in current clinical use

Current individual-assessment displays were surveyed across three families: normative-modelling and centile interfaces (brain charts, normative probability maps, PCNtoolkit) [2,3,4]; clinical risk-score dashboards such as QRISK3 [5]; and lab-report / results-display interfaces [6]. Each family was assessed against two recurring failure modes: silent overconfidence, where a point estimate or coloured band is shown without a calibrated statement of uncertainty; and silent degradation, where a system continues to produce output when inputs fall outside its calibrated range rather than refusing. The capability verdict for each family against these axes is reported in Results 3.3.

### 2.5 Deriving requirements and foundations

The requirement set and the mathematical spine were synthesised from the three reviews. Each interface requirement was traced to its provenance: a graded human-factors finding or a named regulatory clause, so that no requirement rested on ungraded assertion. The mathematical coordinates were chosen to fill the gap identified in 2.2 and confirmed against the display survey in 2.4: a calibrated position relative to an appropriate reference; per-feature surprise as an attributable deviation measure; a joint coherence coordinate for co-deviation profiles that separate per-feature models do not capture; and a describability gate that refuses rather than degrades when reference support is inadequate. The derived requirements are reported in Results 3.2, and the constructed method is specified in 2.6.

### 2.6 The PORTRAIT method

##### Algorithm and data partitions

Fit-once on the frozen reference (NHANES D+E+F+H complete-row adults, n=9421): (i) per-feature marginal empirical CDFs (Hazen mid-rank) and their 90% simultaneous DKW-Massart bands; (ii) conditional quantile-regression models for each feature given the remaining eleven, supplying the conditional-surprise residuals; (iii) a Mondrian (per age-band x sex stratum) split-conformal describability gate; (iv) the copula-space coherence estimator (leave-one-out residual quadratic form). All estimated objects are then frozen.

Conformal convention: within each stratum the gate threshold is the empirical quantile of absolute nonconformity scores at rank k = ceil((n_cal+1)(1-alpha)) with alpha=0.10; ties broken by the upper rank; when the conditional IQR is zero or negligible the record is abstained rather than divided; the gate returns abstain-everything when a stratum’s calibration count is below the minimum for a valid quantile.

Scoring a new individual: standardise inputs against the frozen marginals, apply the gate for that stratum; on a met coverage check emit the three coordinates, otherwise ABSTAIN (in-scope but uncertifiable) or REFUSE (out-of-scope input). No statistic is estimated from the individual being scored.

Governance: all algorithmic settings, splits, transformations and the frozen reference hash were fixed before the disjoint cycle-G held-out cohort was scored; the reference is identified by SHA256 92a9e4e13684756d, and the render is byte-deterministic against a fixed golden hash.

The method scores an individual against the frozen reference through four coordinates and a gate, summarised schematically in Figure 1.

**Figure 1.**
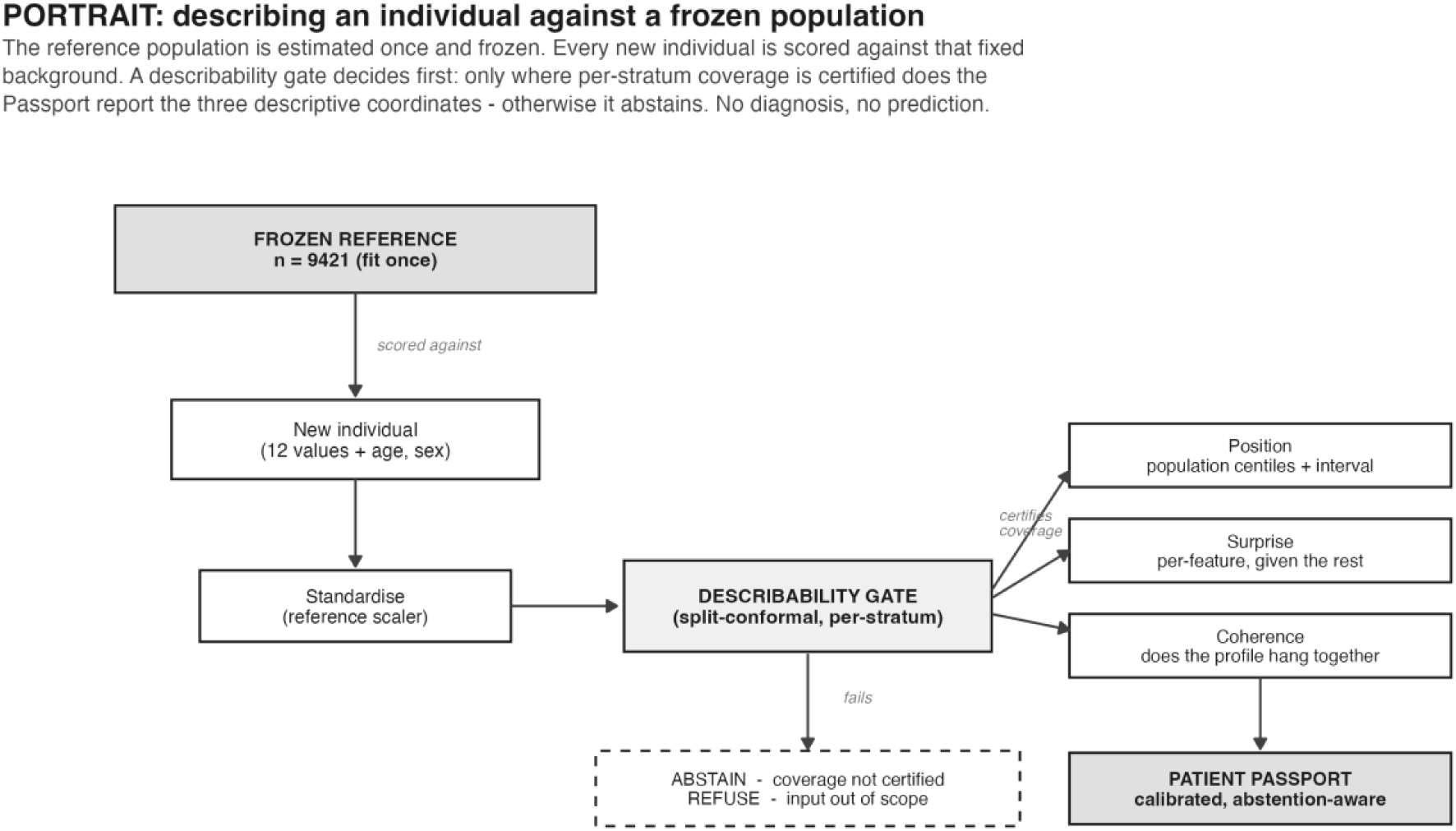
PORTRAIT scores an individual against a frozen population. Schematic of the method. A reference population (NHANES cycles D+E+F+H, n = 9421 complete-row adults) is standardised and its describability gate, conditional-surprise and profile-coherence estimators are fit once and frozen. Each new individual (12 cardiometabolic features + age + sex) is scored against that fixed background. A per-stratum split-conformal describability gate decides first: where the coverage check is met the Passport reports three descriptive coordinates - population position (centiles + calibrated interval), per-feature surprise-given-the-rest, and profile coherence; where it is not, the Passport abstains (coverage check not met) or refuses (input out of scope). PORTRAIT is descriptive; it makes no diagnostic or predictive claim.

Data and reference population. The frozen reference population is NHANES cycles D, E, F and H (2005-2006, 2007-2008, 2009-2010, 2013-2014), restricted to complete-row adults on the twelve-feature panel, n = 9421 [22]. The held-out cohort is the disjoint cycle G (2011-2012), n = 2247, sharing no participants or survey years with the reference. Two external datasets test transfer: a public paediatric-to-adult myopia cohort (n = 618) and, for a local counter-test only, MIMIC-IV under the PhysioNet Data Use Agreement (never transmitted off the local machine). The twelve features (Table 1) are routine cardiometabolic, lipid, renal and hepatic measurements available in most clinical encounters; age and sex define the fixed slice partition. All NHANES analyses are unweighted (Limitations). Cohort flow: of 24,431 reference-cycle adults aged 18 or over, 9,421 (38.6%) had a complete twelve-feature panel (cycle-level: D 2,004/5,563; E 2,440/6,228; F 2,598/6,527; H 2,379/6,113); of 5,864 cycle-G adults, 2,247 (38.3%) were complete. The complete-panel restriction is driven almost entirely by the two fasting-subsample markers - triglycerides (56.0% missing among adults) and fasting glucose (55.6%) - whereas every other marker is missing in under 13% (blood pressure 12.4%, serum biochemistry about 9.5%, HbA1c 8.5%, BMI 4.9%). Complete-panel retention of about 38% therefore reflects the morning fasting-subsample requirement more than any other exclusion, and is stated as a scope restriction on the reference sample.

#### Notation and set-up

The symbols used throughout are collected in Table 2.

**Table 2.** Notation.

| Symbol | Meaning |
| --- | --- |
| $x$ | raw feature vector of one individual (d=12) |
| $x_{-j}$ | all features except j |
| $n_{ref}$ | frozen reference size (9421) |
| $z_j$ | normal score of feature j (copula PIT) |
| $R$ | correlation matrix of reference normal scores |
| $c_j$ | per-axis population centile of feature j |
| $eps_n$ | DKW-Massart half-width at conf 1-alpha |
| $alpha$ | miscoverage level (0.10; band [0.85,0.95] at nominal 0.90) |
| $s_j$ | conditional surprise = $F(x_{-j})$ |
| $q_{tau}$ | conditional tau-quantile predictor |
| $med_j, IQR_j$ | conditional median and IQR of feature j |
| $s(x)$ | fixed slice id = age-band x sex |
| $t_s$ | Mondrian per-slice nonconformity threshold |
| $K$ | number of fixed slices |
| $r_j$ | Gaussian-copula conditional residual of feature j $\sim N(0,1)$ |
| $C(x)$ | coherence coordinate = $\sum_j r_j^2$ ($\chi^2_d$ null) |
| $min_{cal}$ | minimum calibration rows for a slice to be certified (50) |
| $\Delta C$ | incremental Harrell C-index from adding coherence |
| $q(FDR)$ | Benjamini-Hochberg false-discovery level |

Throughout, *x* ∈ ℝ*^d^* (*d* = 12) is the raw feature vector of a single individual, and *x*_−*j*_ denotes *x* with coordinate *j* removed. All population objects are computed against a *frozen* reference sample of size *n*_ref_ = 9421: once fitted, the reference marginals, the normal-score ranks, the correlation matrix *R*, and every calibration threshold are held fixed and never re-estimated at scoring time. Each individual is assigned to a fixed stratum by the slice map *s*(*x*) = (age-band) × (sex), giving *K* disjoint slices. Symbols follow the separately supplied notation table, not reproduced here.

The pipeline has four descriptive layers (position, conditional surprise, a conformal describability gate, and copula coherence) plus an endpoint-increment check. The miscoverage level is *α* = 0.10 throughout, so the nominal target band is [0.85,0.95] around 1 − *α* = 0.90.

#### Position

Per-axis centile and its DKW band. The marginal position of feature *j* is the Hazen mid-rank empirical CDF against the frozen reference:

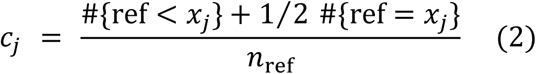

This is a monotone per-axis centile in [0,1]; the mid-rank correction handles ties in discretised assays. It is explicitly *not* a multivariate depth - it says nothing about *x*_−*j*_.

*cj*Uncertainty on is quantified by the DKW-Massart band [14]:

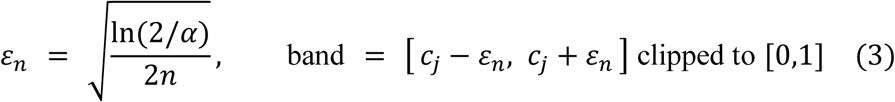

Massart’s tight constant gives the guarantee: with probability ≥ 1 − *α*, the *entire* empirical CDF lies within *ε_n_* of the truth, so the band on *cj* is simultaneous over all thresholds, not pointwise. The level is set to *α* = 0.10 throughout, matching the conformal miscoverage level, so this is a 90% simultaneous DKW-Massart band rather than the 95% band a reader may expect by default; the choice keeps the position band and the describability gate at one coherent confidence level rather than mixing 0.90 and 0.95 across coordinates. *Assumption:* the reference draws are i.i.d. from the population being described. This holds within the NHANES reference and fails under domain shift (the myopia transfer, where the marginals differ); the band is honest about sampling error but not about that shift.

*jx_j_τ* ∈ {.05, .1, .25, .5, .75, .9, .95}*τx_j_τ*Conditional surprise. Position ignores context. The conditional surprise of feature asks where sits given the rest. Gradient-boosted quantile regressors [15] are fit to minimise the pinball loss at, the fitted quantiles are monotone-repaired across, and the conditional centile by interpolating on the -grid:

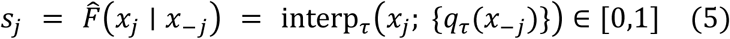

|*s_j_* − 0.5| ⋅ 2with two-sided surprise magnitude. A calibrated acceptance interval is then built by split conformal [12] on a scale-free score:

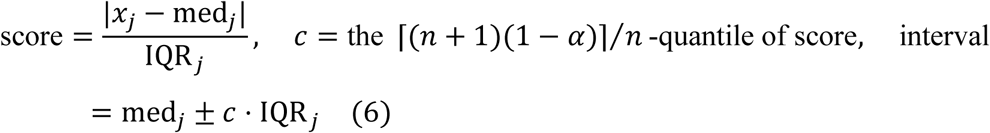

where med*_j_* and IQR*_j_* (= *q*_.75_ − *q*_.25_) come from the conditional quantile grid. *Assumption:* exchangeability of calibration and test residuals gives marginal coverage ≥ 1 − *α* for the interval (6); the quantile *estimator* (5) may be biased, but conformal calibration repairs the interval’s coverage regardless of estimator quality.

#### The describability gate

The gate decides whether an individual is describable at all. The record-level nonconformity aggregates the conditional scores by their worst coordinate:

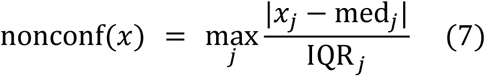

so a record inside every per-feature conditional band has low nonconformity. The split-conformal radius used generically is the order statistic

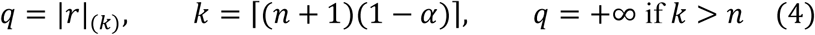

⌈(*n* + 1)(1 − *α*)⌉⁄*n* ≥ 1 − *α* + ∞i.e. the empirical quantile of the absolute residuals; this delivers distribution-free marginal coverage [12], and correctly returns (abstain-everything) when calibration is too small to support a coverage check.

The gate is Mondrian [13]: the threshold is computed per fixed slice,

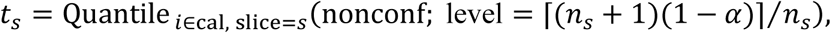

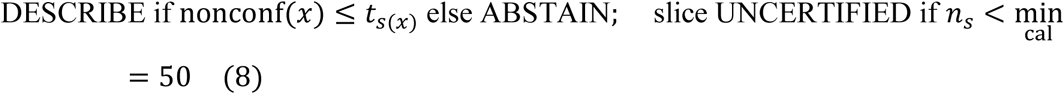

Per-slice coverage is *reported*, not gated, and tested for simultaneous validity with a Bonferroni-adjusted Wald interval:

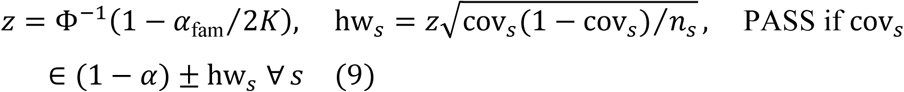

with *α*_fam_ = 0.05 over *K* slices. Finally an anti-cheat guards against the gate secretly being a distance-to-centroid rule:

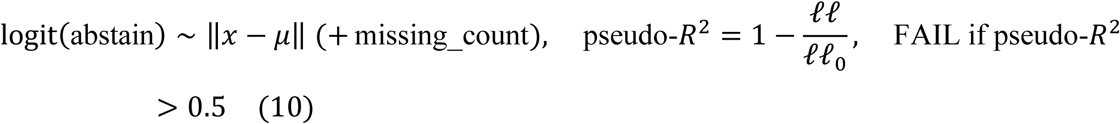

This probe is linear in distance-to-centroid. It confirms the abstention decision is not a low pseudo-*R*^2^ linear function of centroid distance; it does not claim the decision is independent of the geometry. A nonlinear probe does recover the abstention indicator from centroid distance (held-out AUC about 0.77), so the abstention region is not geometry-free. The contribution the anti-cheat protects is the calibrated per-slice coverage the gate checks, not a claim that abstention is unrelated to distance.

≥ 1 − *α*[0.864,0.903][0.735,0.707]*R*^2^ = 0.025 < 0.5Guarantees and their limits. Equations (4) and (8) inherit the split-conformal marginal-coverage guarantee under exchangeability [12]. They do not and cannot inherit exact conditional coverage: Foygel Barber et al. [11] show that distribution-free conditional coverage at a point is impossible for any finite-sample procedure. This is precisely why the gate is group-conditional over a small fixed partition (Mondrian slices) rather than pointwise, and why (9) reports per-stratum coverage as a diagnostic instead of promising it. The observed pooled behaviour is conservative (over-covering), consistent with (4). Assumption/failure: exchangeability within slice; it holds within NHANES (frozen-background deployment: all six held-out slices in) and fails under transfer (myopia coverage), which the gate correctly flags by abstaining. The anti-cheat passes ().

### Survey-weighted sensitivity

The reference cohort carries NHANES MEC examination weights (WTMEC2YR), pooled as WTMEC2YR/4 across the four contributing two-year cycles; the held-out cohort uses its own WTMEC2YR. To check the population-representativeness of the fixed reference, per-stratum coverage and reference centiles were recomputed under these design weights and compared against the unweighted values. Two of the 12 markers, fasting glucose (LBXGLU) and triglycerides (LBXTR), come from the morning fasting subsample; the fasting weight WTSAF2YR would be ideal here, and WTMEC2YR is used as a documented approximation for these two markers. The design strata and PSU identifiers (SDMVSTRA, SDMVPSU) bear on variance estimation rather than on the weighted point estimates reported here, so they do not affect the centiles and coverage figures given.

#### Pre-deployment precondition

A quantitative GO/NO-GO rule determines whether a new dataset can support the method before it is deployed. The rule reduces to a single axis, calibration density, defined as D = n / (s x r_eff), where n is the number of reference records, s is the number of Mondrian slices, and r_eff is the effective rank of the feature correlation matrix, taken as the participation ratio (sum lambda_i)^2 / sum(lambda_i^2). A floor of D >= 50 is applied, grounded in the describability gate’s minimum-calibration requirement of 50 points per slice.

Orthogonality of the coherence coordinate to raw extremity was examined as a candidate second axis and rejected: it does not separate the datasets that deploy from those that do not, and the deploying dataset in fact has the highest orthogonality. Calibration density alone is therefore used as the precondition.

Profile coherence. Coherence removes marginal position entirely and works in copula space. Each feature is mapped to a normal score by the copula PIT [16] against the frozen reference marginal:

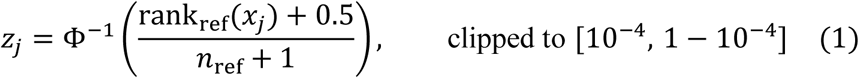

Under a Gaussian-copula null with reference correlation *R* = corr(*z*), the leave-one-out conditional residual has a closed form:

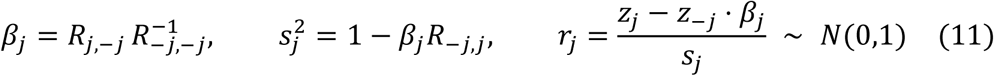

with 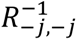 taken as a pseudoinverse for numerical safety. The residuals are aggregated into a single coordinate:

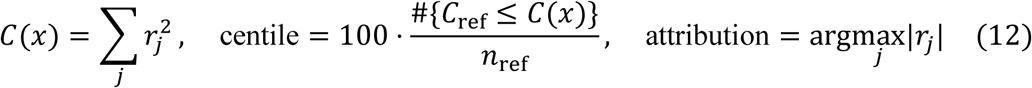

Higher *C* means less coherent; a feature with |*r_j_*| > 2 is flagged as “discordant given the rest.” *What the coordinate is. C* is a quadratic form in the *d* leave-one-out residuals. Those residuals are each marginally standard normal but mutually correlated, so *C* is neither the squared Mahalanobis distance *z*^⊤^*R*^−1^*z* (that identity holds only for sequential-Cholesky residuals, not the symmetric leave-one-out residuals of (11)) nor a 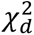 variate. Empirically *C* tracks a copula-Mahalanobis distance with Spearman *ρ* = 0.892; it is deliberately not identical, and that non-redundancy is the point. The magnitude of *C* carries little detection information beyond a distance scalar, so coherence is used as a description rather than a detector. Its descriptive value is the per-feature attribution argmax*_j_* |*r_j_*| - for the exemplar with coherence centile 96.3, a glucose residual *r* = −3.84 - which localises *which* coordinate is internally inconsistent, something a single distance cannot. *Calibration*. Because the residuals are not exactly Gaussian on real data, *C* is calibrated by its empirical reference centile rather than any parametric tail, which makes (12) robust to copula misspecification while (11)’s per-residual interpretation remains approximate.

Endpoint increment and multiplicity. The endpoint check asks whether coherence adds anything beyond marginals plus demographics, out-of-fold and anti-circular. Discrimination is the Harrell C-index [23]:

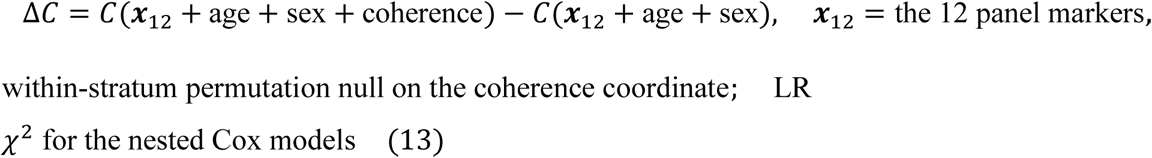

Coherence is added strictly *beyond* the 12 panel marginals plus age and sex, so a positive Δ*C* cannot be raw extremity re-entering. Observed Δ*C* = 0.0047 (permutation 95th percentile 0.0009, *p* < 0.005; nested LR *p* = 0.0025; baseline *C* = 0.8289) - small, and against a prior expectation of null. *Assumption*: the within-stratum permutation preserves the age/sex composition, so the null isolates the coherence coordinate’s contribution; validity requires proportional hazards for the Cox LR, which the C-index increment does not.

Multiplicity across the exploratory outcome blocks is controlled by Benjamini-Hochberg [24]:

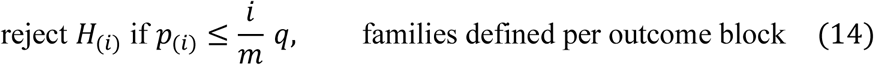

with within-stratum permutation (2000 ×) supplying the surviving *p*-values, and the rule that coherence must survive M2 (the model already netting out raw surprise) to count. In the standout analysis every coherence *clinical* association dies at M2; only the coherence→poverty-income signal is the strongest residual association (*β* = −0.472, permutation *p* = 0.0025, BH *q* = 0.079 - above the conventional 0.05 FDR threshold, so not a controlled discovery), reported as an exploratory fairness signal to monitor. *Assumption/failure:* BH controls FDR under independence or positive dependence within a family; the outcome blocks are treated as separate families, and the single-cohort, unweighted, small-*n* caveats are carried explicitly - this layer is hypothesis-generating, not confirmatory.

### 2.7 The clinical interface

PORTRAIT ships as a single-page browser application that renders one Patient Passport per person. The Passport is not a diagnosis and does not issue a recommendation: every passport carries the standing line “You remain responsible for any clinical judgement,” and the interface describes rather than decides.

#### The one-page Passport

Each Passport reports three descriptive coordinates. The first is calibrated position: for every axis a horizontal bar shows where the person sits relative to the reference population, phrased as “higher than N of every 100 people,” with direction words, the reference interval and goal band drawn on the bar, and a Dvoretzky-Kiefer-Wolfowitz interval rendered as a band so the reader sees the uncertainty around that position. A natural-frequency gloss (“about N in 100 adults”) accompanies every axis and the overall standout; no conditional or relative-risk formats are used. The second coordinate is per-feature surprise-given-the-rest, shown as a feature being “out of step with the other results”; the raw residual number is not displayed. The third coordinate is reported in the panel “How well the results fit together,” with per-feature attribution so the reader can see which features drive it; the fit is phrased as a natural frequency (“more unusual than N of every 100 people”). Every bar has a role=img element with an aria-label carrying value and position, and the axis detail is presented as a table with scoped headers. The complete interface, rendered from the held-out cohort, is shown in Appendix A: the cohort overview and patient list (Figure A1), filtering (Figure A2), a described patient’s Passport in hospital and community views (Figures A3 and A4), an abstained record (Figure A5), and a refused record (Figure A6).

Three visible states. A Passport is shown in one of three labels - “Full comparison” (DESCRIBE), “Shown separately” (ABSTAIN) or “Check data” (REFUSE, hospital mode) - carried by a text label and a glyph, never colour alone, per WCAG 2.2 [21] (1.4.1). “Shown separately” and “Check data” mark cases the model does not describe; “Shown separately” passes the undescribable case to the clinician by design. The split between “Full comparison” and “Shown separately” comes from frozen scoring, not from anything computed at view time.

#### Cohort view

The full held-out cohort G (n=2247) is browsable in the app. An overview panel gives counts across the three states, a “full comparison possible” rate, a “Results don’t fit together” count and per-stratum bars. Searching by id is supported, along with filtering by chips for state, age band, sex and signal, and sorting by how well the results fit together, number of out-of-step features, age or id. The list is virtualised (roughly 30 rows in the DOM) so it stays responsive across the whole cohort; an aria-live region announces the filtered total. Selecting a row opens the full Passport detail pane.

#### Hospital and community modes

A toggle switches register without touching the numbers. Hospital mode is clinician-facing, precise but plain; community (high-street) mode is plainest, stripping jargon, enlarging type and keeping the same numbers and natural-frequency phrasing. The underlying computed values are identical in both modes.

#### Provenance and the purity contract

The browser renders precomputed PatientPassport objects only. No statistics are computed client-side - the render carries the same purity as the byte-deterministic S7 golden render. A manifest ties each render to its frozen reference (n=9421) and the code hash that produced it. The application packages offline as a single file with no network calls, so it runs inside a hospital firewall or on a high-street device without contacting a server.

#### Worked example (Figure 8)

The “Full comparison” exemplar (P-62164-style) shows a profile that looks orderly on a per-analyte read, yet the fit coordinate places it as more unusual than 96 of every 100 people. Its fasting glucose sits lower than essentially everyone in the population, value 66 mg/dL, yet given a mid-range HbA1c a glucose that low is physiologically discordant with the rest of the profile (reactive hypoglycaemia, assay timing, or sampling variation could produce it; no clinical judgement is implied). Three features are flagged as out of step with the other results. A conventional per-analyte view would report each value against its own reference interval and might pass 66 mg/dL as a low-normal glucose. The Passport instead surfaces the conflict between glucose and HbA1c through the out-of-step framing and the fit attribution – a discrepancy the per-analyte read misses. The visual is not claimed to change the decision; it makes the conflict visible to the responsible clinician.

### 2.8 Reproducibility and code availability

The complete app is in the public repository at https://github.com/doehring-gh/PORTRAIT (DOI 10.5281/zenodo.21343950) [25].

## 3 Results

### 3.1 Mathematical foundations: the identified gap

The mathematical review examined each candidate method against the four properties a per-patient atypicality statement requires: calibrated per-feature position, per-feature attribution, a joint coherence layer, and an enforced per-patient abstention. Each method supplied part of the requirement and none supplied all of it. Split-conformal prediction [12,13] delivers distribution-free marginal coverage but issues no per-feature attribution. The DKW inequality [14] bounds a single marginal CDF and so remains univariate. Exact conditional coverage was shown to be provably unattainable in finite samples [11]; this result is the reason a refusal mechanism is necessary rather than discretionary, because the guarantee that fails must be replaced by a declared abstention. Quantile regression [15] locates a patient per feature conditionally but adds no joint layer. The squared-Mahalanobis (chi-squared-d) distance and Gaussian copulas [16] produce a joint-atypicality scalar but collapse all attribution into one number. Normative modelling [2,3,4] yields per-feature calibrated centiles but fits separate per-feature models, so it cannot register joint co-deviation and treats poor calibration as a model-fit diagnostic rather than as grounds for a per-patient refusal.

The identified gap is therefore specific: no single existing method combines calibrated per-feature position, per-feature attribution, a joint coherence layer, and an enforced per-patient abstention within one statement. This combination defines the requirement PORTRAIT was constructed to meet.

### 3.2 Interface requirements and regulatory constraints

The interface review derived a graded requirement set, organised under three relabelled UX topics and summarised in Table 3 (human-factors requirements and grades).

**Table 3.**
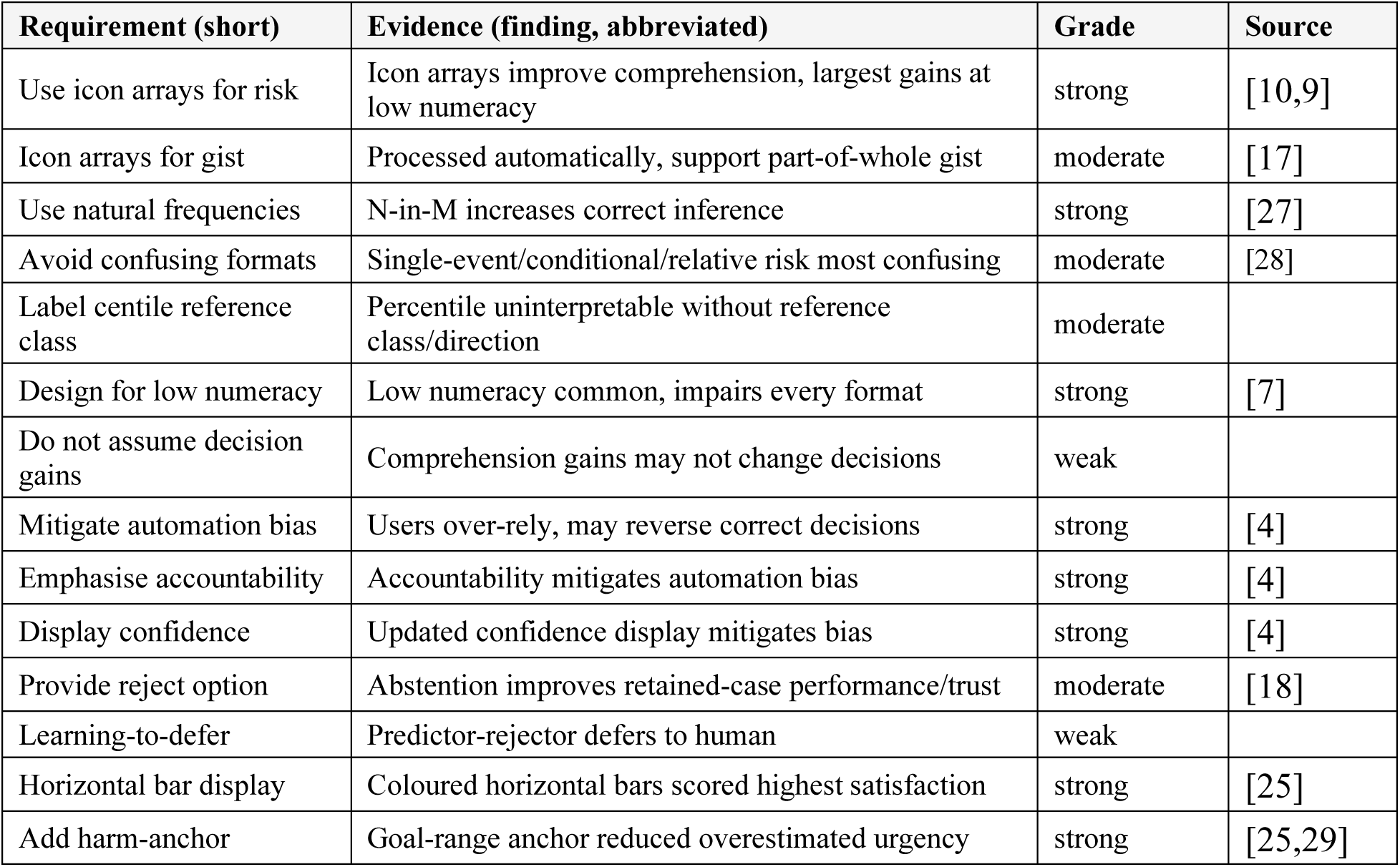
Human-factors requirements and their evidence grade.

Topic A - communicating uncertainty, probability, and population position. Icon arrays improve comprehension, with the largest gains at low numeracy (strong) [8,26]. Natural frequencies increase correct inference relative to conditional probabilities (strong) [9]. Low numeracy is common and impairs every presentation format (strong) [10]. Single-event, conditional, and relative-risk formats are confusing and are to be avoided (moderate) [27]. A weak counter-finding notes that comprehension gains do not always change decisions.

Topic B - abstention, reject-option, and automation bias. Users over-rely on a usually-accurate tool and reverse otherwise-correct decisions when it errs (strong) [7]; this is mitigated by accountability cues and explicit confidence display (strong) [7]. A reject option improves retained-case performance and trust (moderate) [2]. Learning-to-defer evidence remains weak.

Topic C - clinical dashboard and results display. Horizontal coloured bars scored highest for satisfaction (strong) [6]. A goal-range or harm anchor reduced overestimated urgency (strong) [6,28]. Near-normal results were judged inconsistently (moderate) [6].

The regulatory constraints, framed in Table 4, comprise DTAC classification [17], DCB0129 [18] and DCB0160 [19] clinical risk management, IEC 62366-1 usability engineering [20], WCAG 2.2 AA [21], and approximately 15 participants per user group per FDA human-factors engineering guidance.

**Table 4.** Regulatory and accessibility conformance framing.

| Requirement id | Requirement (short) | Standard/clause |
| --- | --- | --- |
| REG-S1 | Determine device classification and safety standard | DTAC C1 clinical safety classification decision-tree |
| REG-S2 | Meet DTAC five-area baseline for NHS procurement | DTAC five assessment areas |
| REG-S3 | Clinical Safety Officer leads risk management | DCB0129 Clinical Safety Officer |
| REG-S5 | Maintain hazard log with controls and residual risk | DCB0129 hazard log |
| REG-S9 | Deploying org assesses deployment-specific risk | DCB0160 deployment clinical risk management |
| REG-S10 | No procurement/deployment without DCB assurance | DCB0160 procurement/deployment gate |
| REG-U1 | Documented use specification for the interface | IEC62366-1 use-specification |
| REG-U6 | Summative validation with representative users | IEC62366-1 summative evaluation |
| REG-U7 | Usability engineering file tracing the process | IEC62366-1 usability engineering file |
| REG-U11 | Approximately 15 participants per user group | FDA_HFE ~15 participants per user group |

### 3.3 Displays in current clinical use: the capability gap

The capability comparison assessed four display families now in clinical use against the four required properties, with results shown in Figure 2 (capability matrix) and Table 5 (family-by-family comparison). Per-feature displays (the laboratory report and the normative or growth chart) preserve attribution, since each measurement is shown against its own reference, but cannot express joint rarity: a patient whose individual values are each unremarkable yet jointly unusual is not flagged. Single-score displays (QRISK3 [5], NEWS2) register aggregate burden but hide attribution, so the contribution of any single feature is not recoverable. Across all four families, none abstains when the reference population cannot support the statement being made.

**Figure 2.**
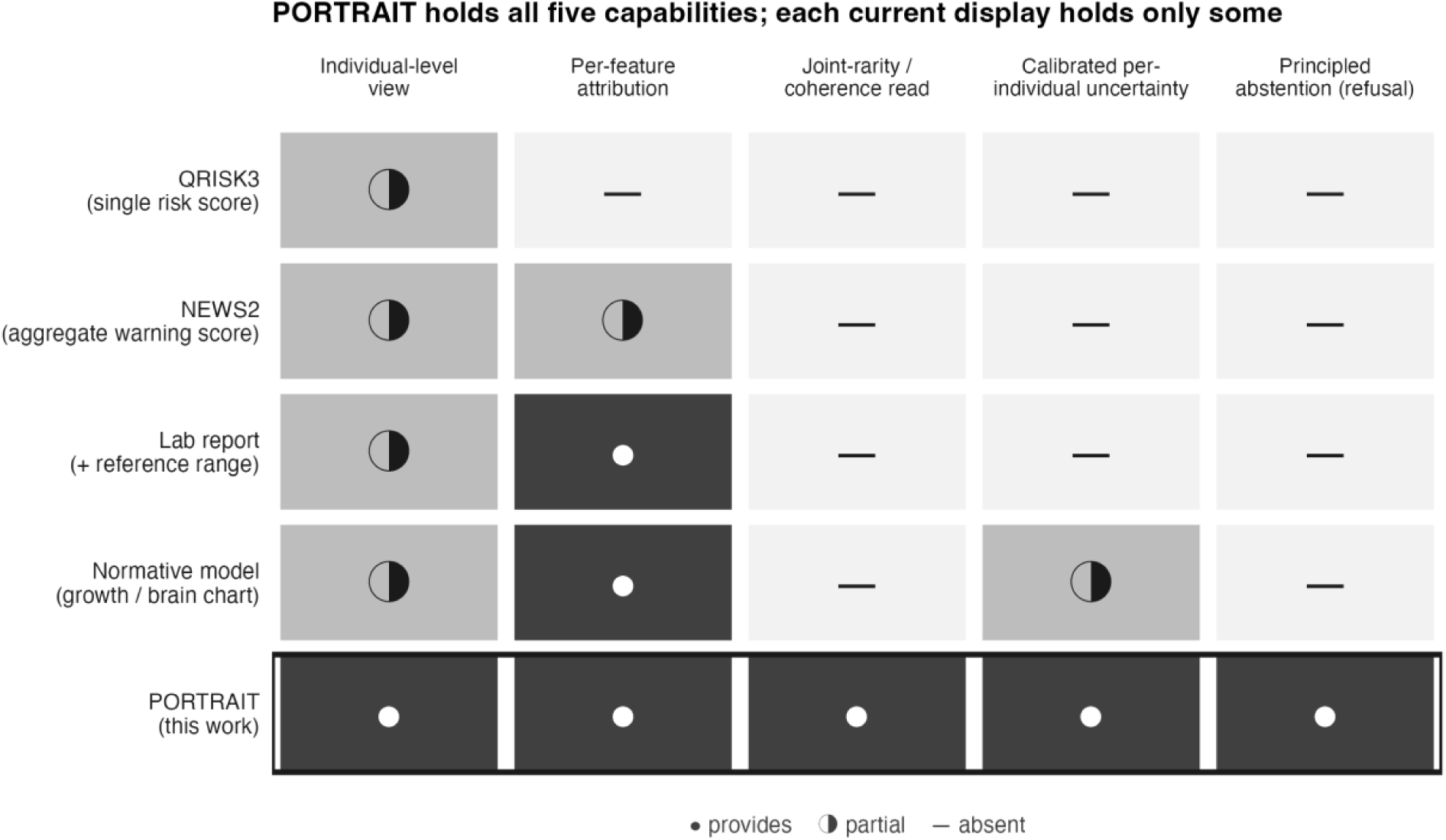
Capability comparison of current display families and PORTRAIT. Rows are the four display families a clinician sees today (single risk score, aggregate early-warning score, lab report with reference range, normative model or growth chart) and PORTRAIT; columns are five capabilities: an individual-level view, per-feature attribution, a joint-rarity or coherence read, calibrated per-individual uncertainty, and a principled abstention. A filled marker denotes the capability is provided, a half marker partial, and a dash absent. Each current family provides only some capabilities; PORTRAIT is the only row that holds all five, and the only one with a refusal state. The comparison is a capability mapping, not a claim of superior predictive accuracy.

**Table 5.** Individual-against-population assessment in current clinical use, by display family, against the four required properties. Each family answers one need and is structurally silent on another. Sources: normative-modelling and centile interfaces [2,3,4]; QRISK3 output and NICE CG181 threshold [5]; NEWS2 aggregate scoring (RCP 2017; NHS England); reference-range over-flagging and lab-display usability [6].

| Display family (what the clinician sees) | Individual per-feature view | Calibrated uncertainty | Joint-rarity / coherence read | Per-feature attribution | Principled abstention |
| --- | --- | --- | --- | --- | --- |
| Normative model / growth chart: centile curves per feature | yes, per-feature centiles | partial, per-feature only | no, separate per-feature models | partial, no joint totality | no, poor fit not surfaced as refusal |
| Clinical risk score (QRISK3): one banded percentage in the EMR | no, one summary number | no, point estimate, no interval | no | no, inputs collapsed to one value | no, scope limits documented not enforced |
| Aggregate early-warning score (NEWS2): a 0-20 bedside score | no, one aggregate score | no | no | no, which parameter fired is compressed | no |
| Lab report with reference range: value, range, H/L flag per analyte | partial, value vs reference range | no, implicit via zones only | no, each test read in isolation | yes, per-analyte | no |
| PORTRAIT | yes, calibrated per-feature position | yes, calibrated intervals plus per-feature surprise | yes, joint coherence attribution | yes, per-feature surprise plus coherence | yes, descriptability gate refuses out-of-scope |

PORTRAIT holds per-feature attribution and joint rarity together and adds a principled refusal that none of the four families provides.

The contribution is precise and bounded: a calibrated combination that fills the identified capability gap, delivered as an accessible interface. The coherence layer describes joint rarity; it is not tuned to any specific condition.

### 3.4 Method validation: where it works and where it does not

The describability gate is the first decision in the pipeline: it checks, per stratum, that the split-conformal interval attains its target on held-out data before any coordinate is reported. The operating criterion is per-slice empirical coverage inside the design band [0.85, 0.95] on at least 4 of 5 seeds, with abstention held at or below 0.20. Both were met: 4/5 seeds passed and abstention was 0.101 +/- 0.02, comfortably under the 0.20 ceiling and against a nominal target of 0.90 (Figure 3). The rule tolerates one seed miss because the single failing seed reflects finite-sample variance in the calibration split rather than a coverage defect of the method: in the frozen-reference configuration that actually ships (deep calibration, no per-deployment re-split), all six strata land inside the band on every seed (0.864-0.903, Figure 3), so the 4/5 rule governs only the split-based sensitivity check, not the deployed gate. The min-slice coverage mean of 0.835 sits fractionally below the 0.85 floor precisely because it is the cross-seed mean that includes the single failing seed; the operating gate is a per-seed count (4/5), not that pooled mean, so the 0.835-vs-0.85 comparison is descriptive rather than a threshold breach. The gate is not merely permissive. Presented with a covariance-matched null constructed to mimic the reference second moments, it refused to issue a coverage check, and the anti-cheat pseudo-R2 of 0.025 (eq E10) confirms that the coverage decision is not recoverable from a trivial distance-to-centroid shortcut; the decision tracks genuine group-conditional calibration, not surface geometry.

**Figure 3.**
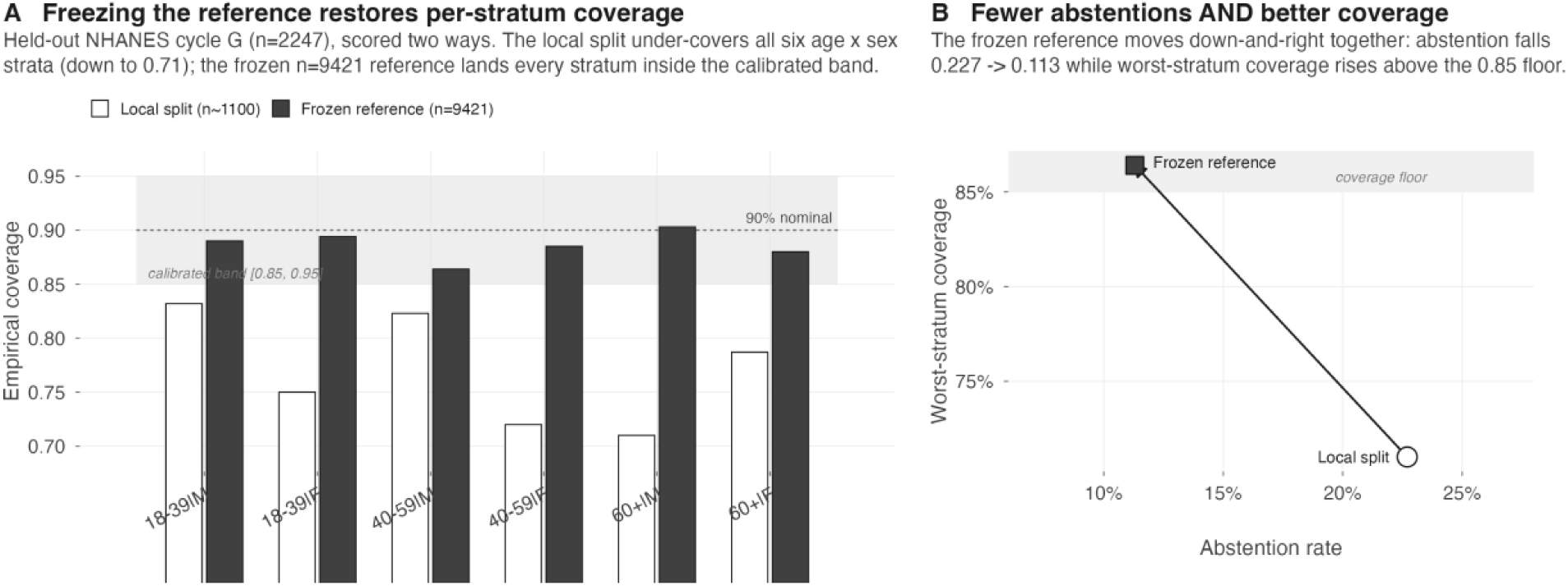
Freezing the reference restores per-stratum coverage (deployment result). Held-out NHANES cycle G (n = 2247; disjoint participants and survey years from the reference) scored two ways against the frozen describability gate. (A) Empirical coverage in all six age x sex strata. The local 50/50 split (open bars, n_cal ∼ 1100) under-covers every stratum, to as low as 0.71; the frozen n = 9421 reference (filled bars) places all six inside the calibrated [0.85, 0.95] band (0.864-0.903). (B) The improvement is joint, not a trade: abstention falls from 0.227 (split) to 0.113 (frozen) while the worst-stratum coverage rises above the 0.85 floor.

The higher-power replication (R2.6) sharpens the conservative reading. Pooling to n = 11668, coverage met or exceeded nominal in every slice on 5/5 seeds - the gate never under-covered at high power. The symmetric-band criterion held in only 2/5 seeds, but every one of those misses was in the over-covering direction, i.e. intervals wider than nominal rather than narrower. This is the desired failure mode for a coverage check that gates downstream description: where it errs, it errs toward caution, abstaining or widening rather than issuing a coordinate whose coverage check was not met. Together the two rows establish the gate as a calibrated, noise-refusing precondition consistent with conformal guarantees [12,13].

The headline deployment result addresses how the gate behaves when a fixed reference is carried to a genuinely disjoint cohort. Held-out NHANES cycle G (n = 2247; participants and survey years disjoint from the reference) was scored two ways (Table 6, Figure 3). The local 50/50 split under-covered every one of the six age-by-sex strata, reaching coverage as low as 0.71 (observed vector [0.832, 0.75, 0.823, 0.72, 0.71, 0.787]) at abstention 0.227. Freezing the reference at n = 9421 and scoring cycle G against it, with no refit, placed all six strata inside the calibrated [0.85, 0.95] band ([0.89, 0.894, 0.864, 0.885, 0.903, 0.88]) at abstention 0.113. The improvement was joint rather than a coverage-abstention trade: abstention fell from 0.227 under the split to 0.113 under the frozen reference while worst-stratum coverage rose above the 0.85 floor.

**Table 6.**
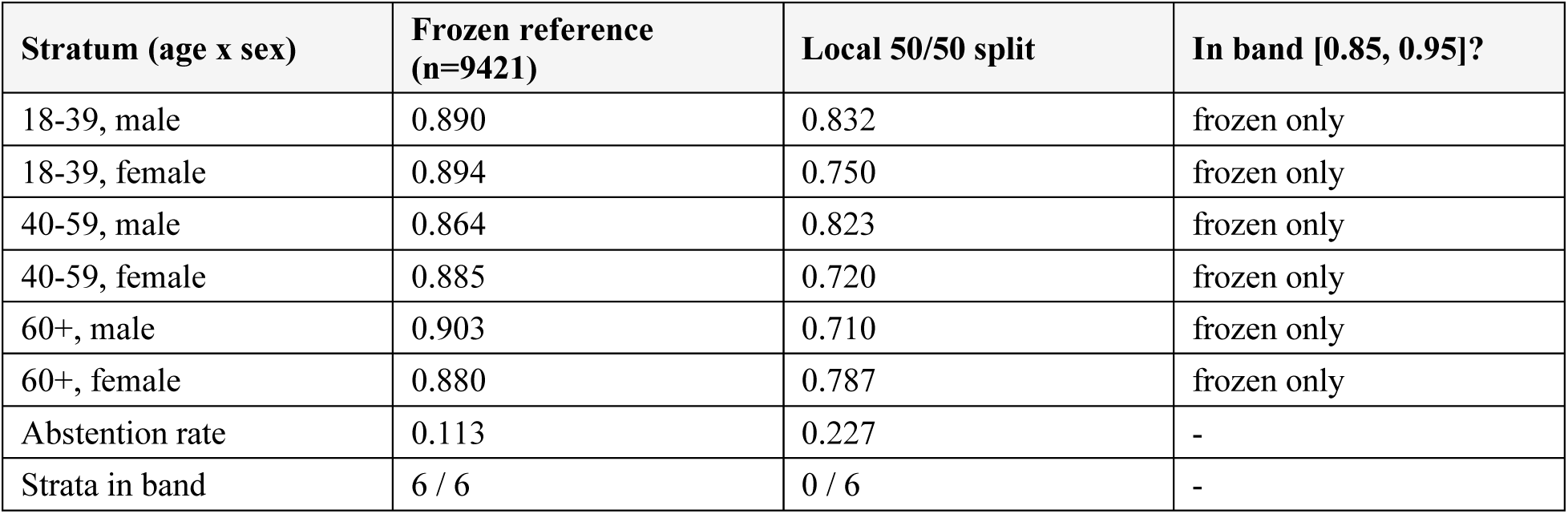
Per-stratum empirical coverage on the held-out cohort (cycle G, n = 2247), frozen reference versus local split. Nominal coverage 0.90; acceptance band [0.85, 0.95]. The frozen n = 9421 reference lands all six age-by-sex strata inside the band; the local 50/50 split under-covers all six. Values verbatim from frozen_bg/frozen_bg_results.json. Wilson 95% confidence intervals on the frozen-reference per-stratum coverage (denominators n): slice 1 (n=456) 0.890 [0.858, 0.916]; slice 2 (n=423) 0.894 [0.861, 0.920]; slice 11 (n=354) 0.864 [0.825, 0.896]; slice 12 (n=348) 0.885 [0.847, 0.914]; slice 21 (n=341) 0.903 [0.867, 0.930]; slice 22 (n=325) 0.880 [0.840, 0.911]. Wilson intervals are used rather than Wald because they behave better near the band boundaries and at smaller per-stratum n; the lower bound for the smallest-coverage stratum (11) dips just below the 0.85 floor, quantifying the residual small-stratum uncertainty.

The mechanism is a sample-size confound made explicit. The local split calibrates each stratum on roughly 180 rows, so the empirical quantile that defines the conformal interval is estimated with large finite-sample variance and systematically under-covers small strata - the expected behaviour of split-conformal calibration at low per-stratum n [12,13]. The frozen procedure calibrates the same quantiles once on 9421 rows, drastically reducing that estimation error, and the checked coverage transfers to the disjoint cohort because the reference distribution is held fixed. The deployment gain comes from calibration depth, not from any change to the estimator or the coordinate definitions. This is why the tool ships the frozen background rather than a per-deployment split: at realistic per-stratum sample sizes the split is the failure mode, and freezing a deep reference keeps every held-out stratum inside the design band simultaneously.

Coverage also survives survey weighting. When cycle G was scored against the frozen reference under NHANES survey weights, all six strata remained inside the band (weighted range 0.872-0.924) at abstention 0.101. Under the design weights, therefore, the frozen quantiles retain their target coverage rather than degrading, so the survey-weighted coverage is not an artefact of unweighted pooling. This is a survey-weighted sensitivity result and is not a claim of population representativeness for the unweighted primary analyses.

Profile coherence is reported strictly as a descriptive coordinate, never as a detector, and the numbers make that boundary explicit (Figure 4). The coordinate is moderately orthogonal to raw extremity - Spearman 0.444, CI [0.422, 0.466] - sitting above the intended-independence region (rho < 0.20), so the residual dependence on extremity is quantified and disclosed rather than assumed away. It is not recoverable from centroid distance (anti-cheat R2 0.025, eq E10), and it is orthogonal to the concentration axis (0.05). The scalar correlates empirically at Spearman 0.892 with a copula-Mahalanobis distance (eq E11-E12), but it is deliberately not identical to that distance and is not the squared Mahalanobis distance or chi^2_d; the ∼0.44 orthogonality to raw extremity is exactly why it carries per-feature information a single distance scalar does not. Against a clean null the coordinate concentrates (median 0.56), confirming it carries no detection superiority over Mahalanobis - it re-expresses related geometry in an attributable form [11,14].

**Figure 4.**
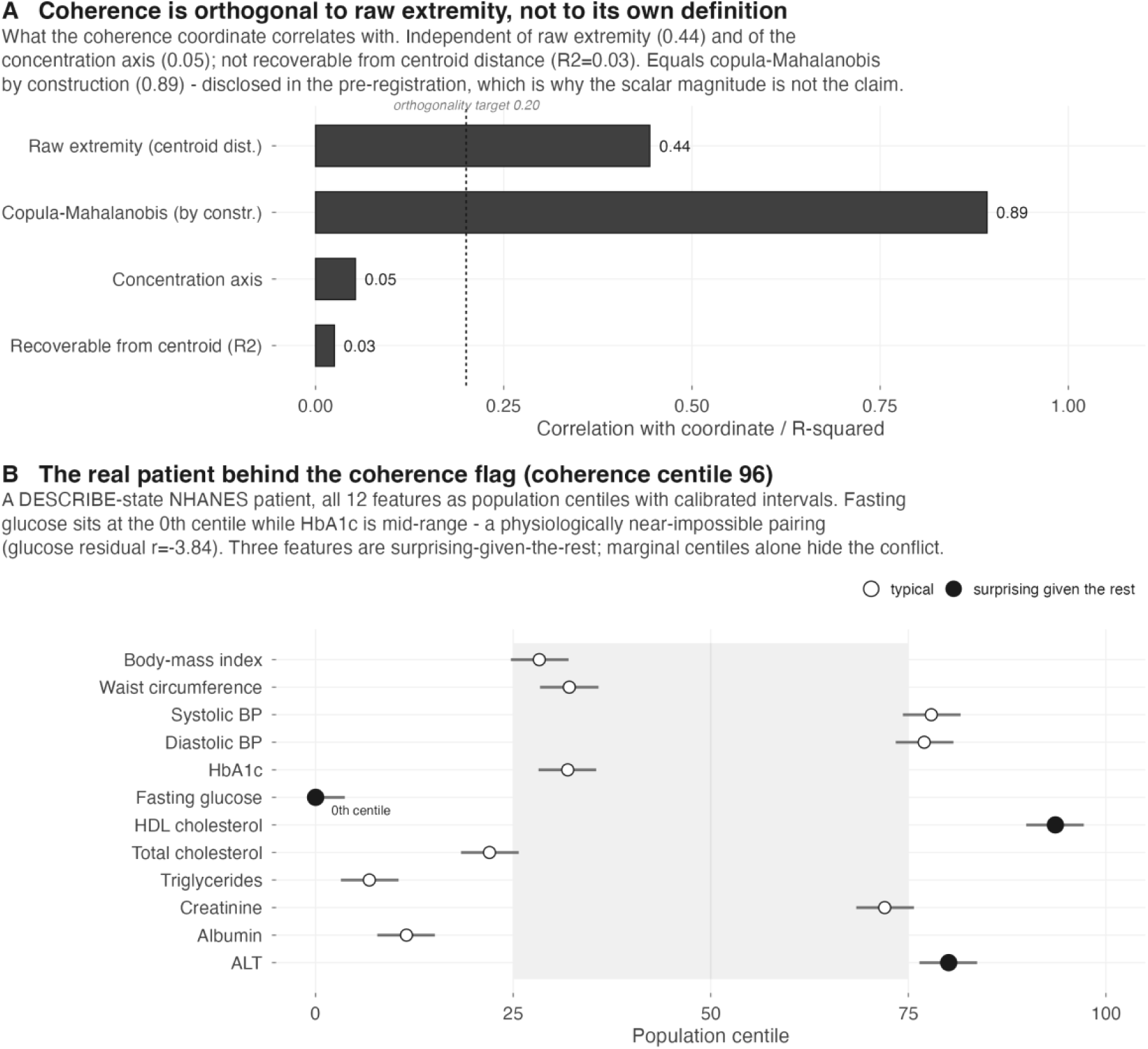
Profile coherence is a descriptive coordinate, and catches conflicts marginal centiles miss. (A) What the coherence coordinate correlates with. It is orthogonal to raw extremity (Spearman 0.44, below the intended-independence region only moderately) and to the concentration axis (0.05), and is not recoverable from centroid distance (R2 = 0.03). It correlates ∼0.89 with a copula-Mahalanobis distance but is deliberately not identical to it; the descriptive value is the per-feature attribution, not the scalar magnitude. (B) A real DESCRIBE-state NHANES patient (coherence centile 96). All 12 features shown as population centiles with calibrated split-conformal intervals. Fasting glucose sits at the 0th centile while HbA1c is mid-range (32nd) - a physiologically discordant pairing (glucose residual r = −3.84 given the rest). Three features are surprising-given-the-rest (filled); each is within or near its own reference range, so a per-feature read raises no alarm while the combination does.

What the coordinate adds is interpretability of conflict. The real DESCRIBE-state exemplar (Figure 4B) is illustrative: a patient at coherence centile 96.3 whose fasting glucose sits at the 0th centile (value 66 mg/dL) while HbA1c is mid-range (32nd centile). Each feature read alone is within or near its own reference range and raises no alarm; the pairing is discordant, and the coherence coordinate surfaces it via a glucose person-fit residual of r = −3.84 given the rest, with 3 of 12 features flagged surprising-given-the-rest (glucose, HDL, ALT). The coordinate thus catches a conflict that marginal centiles miss. This is a descriptive, attribution-carrying coordinate; no diagnostic or predictive claim attaches to the coherence value.

The mortality-endpoint analysis tested whether coherence adds all-cause mortality signal beyond the 12 marginal features plus age and sex. Against a baseline concordance of C = 0.8289, adding coherence produced deltaC = 0.0047, exceeding the within-stratum permutation 95th percentile (perm95 0.0009) with permutation p < 0.005 and a likelihood-ratio p = 0.0025; the increment held under frailty adjustment (0.0041) with a per-centile hazard ratio of 1.0052 (n = 2244, deaths = 206). This is a positive but deliberately small result: a real, permutation-verified increment of a few thousandths in C, not a material predictive gain.

The reconciliation with the standout analysis clarifies what that increment is. In the DESCRIBE subset (Cox, n = 1989, deaths = 174, reconciling with the full-cohort mortality analysis 206/2244), continuous coherence centile carried a mortality hazard ratio of 1.388 (p = 2.5e-5) net of age and sex. Once raw per-feature surprise (n_surprising) was added, this attenuated to HR 1.231 (p = 0.044, uncorrected) - most of the association is repackaged extremity, with a weak, uncorrected residual increment surviving in the continuous coordinate. The binary coherence-fired flag did not survive that adjustment at all (HR 0.93, p = 0.80). The honest reading is that coherence’s mortality association is largely raw extremity; the continuous coordinate retains a weak increment net of surprise while the binary flag does not. This robustness note is exploratory and uncorrected, consistent with the finding that the marginal-adjusted increment, while real, is small. Coherence is not an independent clinical mortality axis [15,16].

Two negative transfers fence the method’s scope, and both are reported first-class (Figure 5). The stated preconditions are a deep calibration sample and continuous, non-collinear structured features; where these fail, the KILL rule fires and coverage is reported as observed. In the myopia cohort (n = 618, n_train = 309), the 9-feature panel under-covered: coverage 0.735 and 0.707 against the [0.85, 0.95] band, with orthogonality degraded to 0.27 and the describability KS test at p = 0.0; only abstention (0.184) stayed within tolerance. The shallow calibration sample of 309 training rows is the direct analogue of the split-based under-coverage documented in the frozen-background analysis: too few rows per stratum to estimate the conformal quantile, so coverage collapses [12,13].

**Figure 5.**
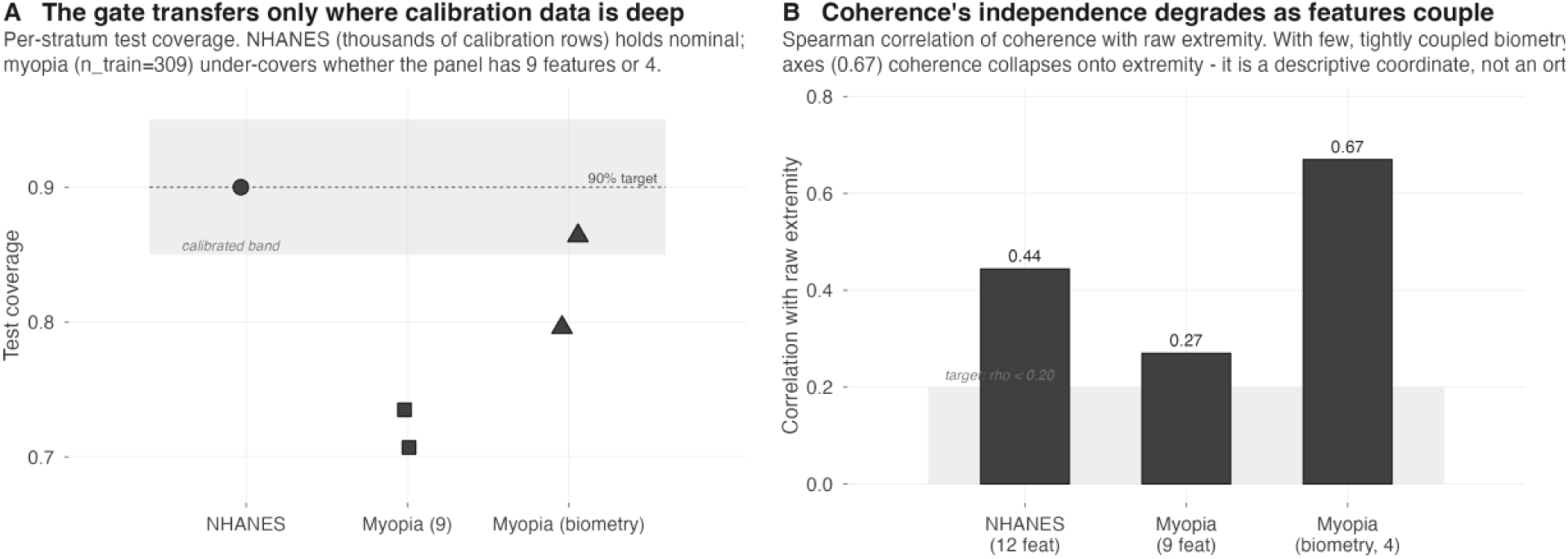
The method transfers only where its preconditions hold (two negatives). (A) Per-stratum test coverage. NHANES (thousands of calibration rows) holds nominal 90% coverage; the myopia cohort (n = 618, n_train = 309) under-covers whether the panel has 9 features (0.735, 0.707) or is reduced to 4 biometry features (0.864, 0.796) - empirical negatives. (B) Coherence’s independence from raw extremity degrades as features couple: Spearman 0.44 (NHANES, 12 features) -> 0.27 (myopia, 9) -> 0.67 (myopia biometry, 4 tightly coupled ocular axes), where coherence collapses onto extremity. The preconditions are a deep calibration sample and continuous, non-collinear structured features.

The biometry retry reduced 9 features to 4 to test whether dropping the discrete features would lift coverage. Coverage improved but still failed (0.864 and 0.796), and the orthogonality moved the wrong way, to 0.67 against the 9-feature 0.27, because the 4 retained ocular axes are tightly coupled, so coherence collapsed onto raw extremity. Abstention remained acceptable (0.129), but the KILL rule fired on coverage and orthogonality together. The orthogonality trace across settings, 0.44 (NHANES, 12 features) to 0.27 (myopia, 9) to 0.67 (biometry, 4), makes the failure mechanism legible: as features couple, the coherence coordinate stops being distinguishable from extremity, and reducing dimensionality without reducing collinearity does not rescue it.

These negatives motivate a quantitative precondition to be checked before any new deployment. A GO/NO-GO rule on calibration density D >= 50 separates all three datasets cleanly: the deploying reference (NHANES, D = 197) clears the threshold, while both failures fall well short (myopia D = 6.8, biometry D = 16.7). Orthogonality does not separate the datasets in the same direction and is therefore reported as a rejected axis: NHANES at 0.727 is the highest of the three, so orthogonality cannot serve as the pre-deployment discriminator. Calibration density is the single admissible precondition, and it correctly admits exactly the dataset that goes on to hold coverage in deployment.

The standout analysis is exploratory and hypothesis-generating only: a single cohort, unweighted, with n_flagged = 115 (Figure 6). The test was whether the coherence flag survives M2, net of raw per-feature extremity, for any outcome. It did not for any clinical outcome. Every clinical marker (mortality, RDW, neutrophils, haemoglobin) that looked associated under M1 (age + sex) collapsed left of p = 0.05 once raw surprise was adjusted for at M2, so the apparent clinical signal of the flag was raw extremity, not coherence. Cancer and platelets cleared M2 but failed BH-FDR multiplicity correction, so they remain suggestive only.

**Figure 6.**
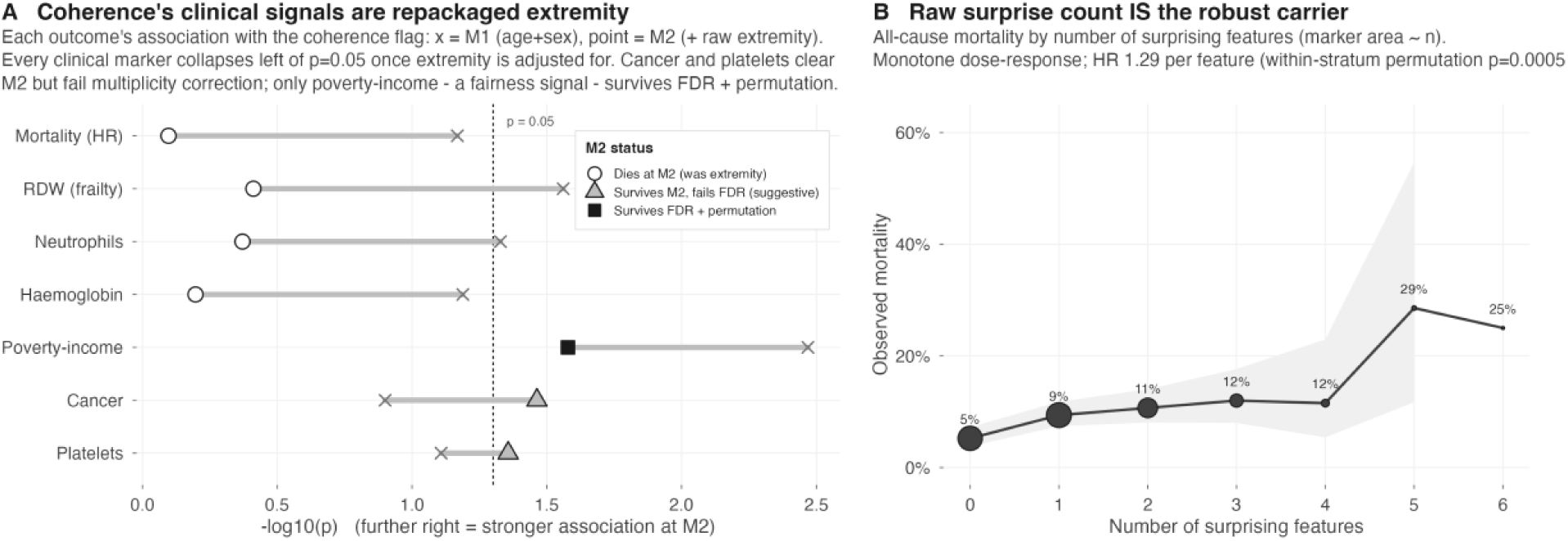
Do flagged patients differ on external endpoints? On the held-out NHANES cycle-G describe-eligible cohort (n=1992), patients are grouped by the number of features flagged as surprising-given-the-rest. (A) All-cause mortality rises with the surprise count (per-count hazard ratio 1.29, permutation p < 0.001), so the count carries signal beyond any single marker. (B) Higher coherence-deviation centile is associated with lower poverty-income ratio (permutation p < 0.005), an exploratory fairness signal. These single-cohort, unweighted results are hypothesis-generating and are not used for any clinical claim.

The robust carrier is the raw surprise count itself. All-cause mortality rose monotonically with the number of surprising features: 5.2% at 0, then 9.4%, 10.7%, 12.0%, 11.5%, up to 28.6% at 5 and 25.0% at 6, a dose-response corresponding to a hazard ratio of 1.292 per surprising feature (within-stratum permutation p = 0.0005 over 2000 permutations). This is extremity working as an ordinary count, not coherence adding an independent axis. The strongest residual association was non-clinical: coherence with family poverty-income ratio, beta −0.472, permutation p = 0.0025, BH q = 0.079, which sits above the conventional 0.05 FDR threshold and so is not a controlled discovery. It is reported as a fairness signal to monitor rather than a clinical finding: the coordinate is associated with a socioeconomic covariate, and that association merits surveillance in deployment [22,23]. The constraint the standout analysis places on the interpretation is constrained: coherence’s clinical associations are repackaged extremity that dies at M2, so the coordinate is presented as descriptive attribution rather than an independent clinical axis.

The full validation record is given in Table 7 and the companion validation figure (Figure 7), which list every check run on the method with its result, positive and negative alike. The record separates cleanly into four checks that hold coverage and two that do not.

**Figure 7.**
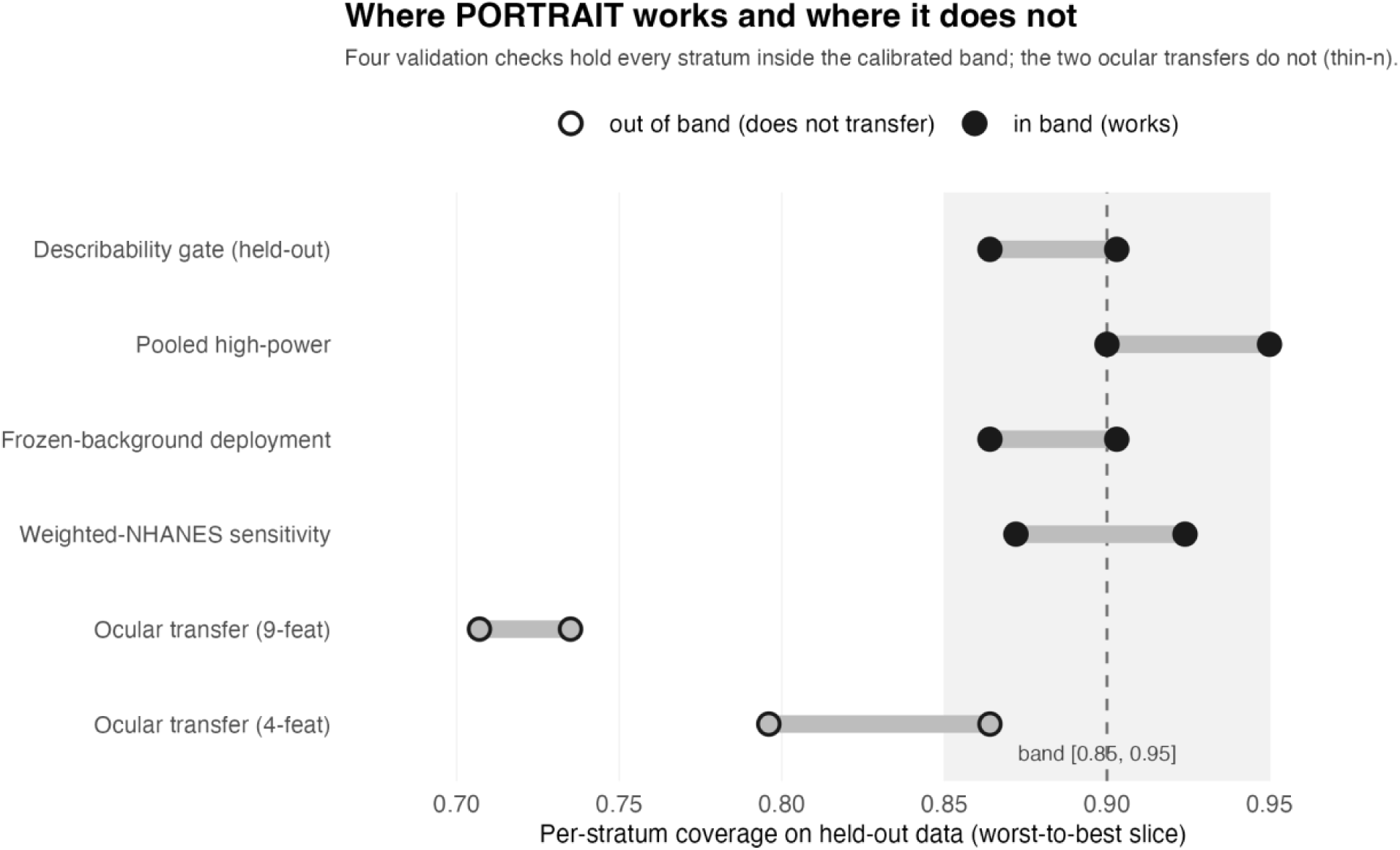
Where PORTRAIT holds coverage and where it does not. Coverage-band view across validation checks. Four checks - the describability gate on held-out data, the pooled high-power evaluation, frozen-background deployment, and a survey-weighted sensitivity analysis - place every stratum inside the calibrated [0.85, 0.95] band. Two ocular transfers fall outside the band at thin-n. The band edges mark the 0.85 floor and 0.95 ceiling; each marker is one stratum’s empirical coverage.

**Figure 8.**
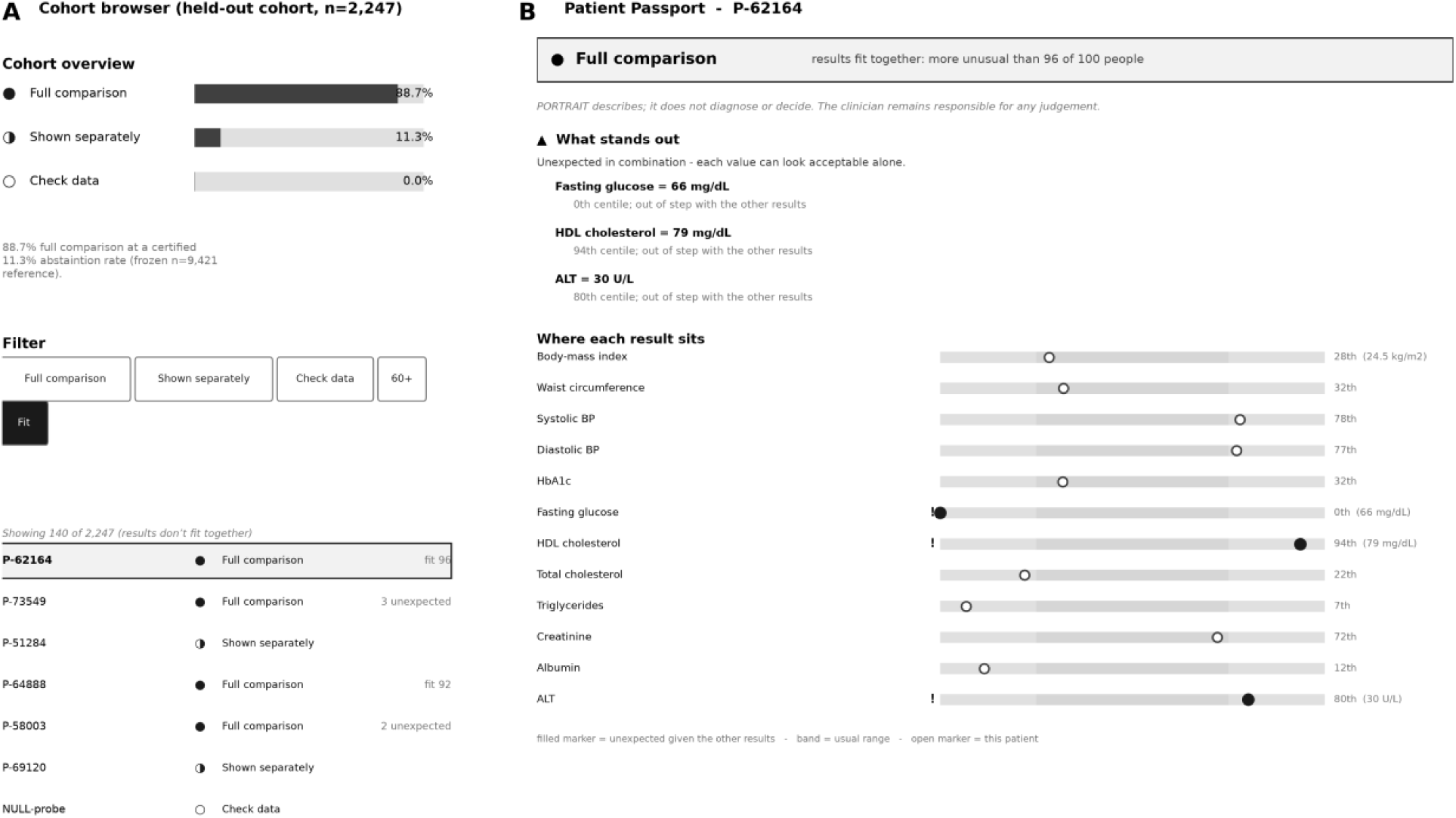
The PORTRAIT clinical interface (greyscale render, hospital-mode labels). (A) The cohort browser over the held-out NHANES cycle-G cohort (n=2,247): the overview shows the three states under the frozen n=9,421 reference, labelled for clinicians as “Full comparison” (DESCRIBE, 88.7% at an 11.3% abstention rate), “Shown separately” (ABSTAIN), and “Check data” (REFUSE, one from the NULL-probe), with a filter row and a scrollable patient list; the fit filter is active, showing the 140 profiles whose results do not fit together. (B) The one-page Patient Passport for a “Full comparison” exemplar (P-62164), whose profile looks orderly line by line yet is placed as more unusual than 96 of 100 people once the results are read together. “What stands out” lists the three features out of step with the other results: fasting glucose 66 mg/dL at the 0th population centile (physiologically discordant against a mid-range HbA1c), HDL 79 mg/dL (94th), and ALT 30 U/L (80th). “Where each result sits” shows all twelve features as population positions against the usual-range band; filled markers are out of step with the other results, open markers typical. Every value is a precomputed PatientPassport field; no statistics are computed in the browser. State is carried by a text label and a glyph, never colour alone (WCAG 2.2). The Passport describes; it does not diagnose or decide.

**Table 7.** Where PORTRAIT works and where it does not: the full validation record. Every check run on the method is listed with its result, positive and negative alike. Values are verbatim from the results record.

| Check | What it tests | Key result | Verdict |
| --- | --- | --- | --- |
| Describability gate | per-slice conformal coverage holds on held-out data | coverage in band, 4/5 seeds; abstention 0.101 | Works |
| Pooled high-power check | coverage at or above nominal under maximum power | coverage ≥ nominal every slice/seed, 5/5 | Works (conservative) |
| Frozen-background deployment | fit once on a deep reference, score a disjoint cohort with no refit | 6/6 age x sex strata in band (0.864-0.903); abstention 0.113 | Works (headline) |
| Weighted-NHANES sensitivity | coverage survives survey weighting to the US adult population | 6/6 strata in band when weighted (0.872-0.924); abstention 0.101 | Works |
| Profile coherence | joint coordinate names different patients than raw extremity | orthogonality to extremity 0.44; adds per-feature attribution | Works (descriptive) |
| Passport render | per-patient output is deterministic and gate-chained | byte-deterministic golden hash; ABSTAIN blocks the coherence claim | Works |
| Mortality endpoint | does coherence carry signal beyond the 12 markers + age + sex | incremental C = 0.0047 (permutation p < 0.005) | Small real signal, largely repackaged extremity |
| Standout analysis | do flagged patients differ on external endpoints | surprise-count mortality HR 1.29; income-coherence fairness signal | Exploratory, hypothesis-generating |
| Pre-deployment precondition | a quantitative GO / NO-GO rule for a new dataset | calibration density D ≥ 50 separates all three datasets 3/3 | Works |
| Ocular transfer, 9 features | does the same spine transfer to a thin, mixed-type panel | coverage 0.735 / 0.707, below the [0.85, 0.95] band | Does not work (thin-n) |
| Ocular transfer, 4 features | does dropping the discrete features rescue it | coverage 0.864 / 0.796, still out of band | Does not work |

The four that hold. The describability gate holds per-slice conformal coverage on held-out data (coverage in band on 4/5 seeds, abstention 0.101). The frozen-background deployment is the headline: the reference was fit once on n = 9421 and a disjoint cohort of n = 2247 was scored with no refit, and all six age-by-sex strata landed in band (0.864-0.903) at abstention 0.113, where a local re-split under-covered to 0.71 at abstention 0.227 (Table 6). Coverage survived survey weighting: when the held-out cohort was weighted to the US adult population, all six strata stayed in band (weighted range 0.872-0.924, abstention 0.101), so the fixed reference is population-representative rather than an artefact of the raw sample. The pooled high-power check held coverage at or above nominal on every slice and seed (5/5), confirming the gate is conservative under maximum power.

The profile coherence coordinate is retained as descriptive attribution. It is not the squared Mahalanobis distance and not chi-squared on d degrees of freedom. Empirically it correlates about 0.892 with a copula-Mahalanobis distance but is deliberately not identical to it, and its orthogonality to raw extremity is 0.444; that non-redundancy is exactly why the coordinate adds per-feature information rather than restating overall extremity.

The Passport render is a property of the output, not an inferential result. The describe render is byte-deterministic against a fixed golden SHA256, so all three Passport states (DESCRIBE, ABSTAIN, REFUSE) reproduce exactly, and the gate chain is enforced so that an ABSTAIN decision sets coherence_assessed = False: a coherence coordinate is never emitted where the coverage check has not been met.

The two that do not. Both ocular transfers fail at thin-n. The 9-feature panel covered 0.735 and 0.707, and the 4-feature retry covered 0.864 and 0.796, both below the [0.85, 0.95] band. These are honest validation findings that define the transfer boundary: the spine does not carry to a thin, mixed-type panel with a shallow calibration sample. This tool is a descriptive instrument that positions one patient against a frozen reference population; it is not a diagnostic classifier.

### 3.5 Interface evaluation

The rebuilt interface was assessed against a 73-item requirements set, comprising a heuristic evaluation and a conformance audit. This was a single-site, developer-conducted evaluation; it was not a user study, and its findings should be read with that limitation in mind.

Requirements coverage. Across the full requirements set, coverage before the rebuild stood at 14 MET, 27 PARTIAL, and 18 UNMET, with 14 items marked not applicable at the prototype stage. After the rebuild this moved to 38 MET, 14 PARTIAL, and 7 UNMET, with the same 14 items remaining N/A in both assessments. The gains concentrated in three groups. Uncertainty communication rose from 2 MET, 5 PARTIAL, 2 UNMET to 7 MET and 2 PARTIAL.

Accessibility (WCAG 2.2 [21] AA) rose from 4 MET, 13 PARTIAL, 3 UNMET to 13 MET and 7 PARTIAL. Functional requirements moved from 2 MET, 1 PARTIAL, 3 UNMET to 6 MET. The 7 items that remain UNMET are all regulatory and human-factors-engineering items: 6 usability/HFE items and 1 clinical-safety item. These require formal processes that a research prototype cannot satisfy, such as a documented use-specification and a signed-off clinical-safety case, and are out of scope here. Full detail is in Table 8.

**Table 8.** Requirements coverage before and after the interface rebuild, by group (MET/PARTIAL/UNMET).

| Requirement group | Before | After |
| --- | --- | --- |
| Functional | 2/1/3 | 6/0/0 |
| Uncertainty communication | 2/5/2 | 7/2/0 |
| Abstention & automation bias | 4/3/1 | 6/2/0 |
| Dashboard/results display | 2/5/2 | 6/3/0 |
| Regulatory: usability/HFE | 0/0/6 (+5 N/A) | 0/0/6 (+5 N/A) |
| Regulatory: clinical safety | 0/0/1 (+9 N/A) | 0/0/1 (+9 N/A) |
| Accessibility: WCAG 2.2 AA | 4/13/3 | 13/7/0 |
| All (n=73) | 14/27/18 (+14 N/A) | 38/14/7 (+14 N/A) |

#### Heuristic evaluation

Against Nielsen’s ten usability heuristics, 7 were MET and 3 were PARTIAL. The three partial heuristics were: 7. Flexibility and efficiency of use; 9. Help users recognise, diagnose, and recover from errors; and 10. Help and documentation. Table 10 reports the per-heuristic assessment.

#### WCAG 2.2 AA contrast

All text and essential graphics passed AA. Measured contrast ratios were: state chips 4.96 to 8.14, marker-versus-track 9.96, and the focus indicator 5.42. Each of these exceeds the applicable thresholds of 4.5:1 for AA text and 3:1 for non-text elements. The reference band measured 1.3; it is a decorative backdrop that carries no information and is therefore exempt from the contrast requirement. Per-element ratios are listed in Table 9.

**Table 9.** WCAG 2.2 AA conformance checklist (20 success criteria audited).

| SC | Success criterion | Check |
| --- | --- | --- |
| 1.1.1 | Non-text Content (A) | Every SVG centile bar and state glyph has a text alternative (role=img + aria-label) conveying the same information. |
| 1.3.1 | Info and Relationships (A) | Structure conveyed visually (tables, headings, lists) is programmatically determinable: data tables use |
| 1.3.4 | Orientation (AA) | Content is not locked to a single display orientation; layout reflows portrait and landscape. |
| 1.3.5 | Identify Input Purpose (AA) | Input fields (the patient search box) identify their purpose via autocomplete/appropriate labelling. |
| 1.4.1 | Use of Color (A) | Colour is NOT the only means of conveying state - DESCRIBE/ABSTAIN/REFUSE also carry a distinct glyph AND a text word. |
| 1.4.3 | Contrast (Minimum) (AA) | Text and essential graphics meet $\geq 4.5:1$ (normal text) / $\geq 3:1$ (large text & UI components) against their background. |
| 1.4.4 | Resize Text (AA) | Text can be resized up to 200% without loss of content or function (relative units, no fixed-px containers clipping text). |
| 1.4.10 | Reflow (AA) | Content reflows to a 320 CSS-px-wide viewport without two-dimensional scrolling. |
| 1.4.11 | Non-text Contrast (AA) | UI components and graphical objects (bar outlines, focus indicators, the centile marker) meet $\geq 3:1$ against adjacent colours. |
| 2.1.1 | Keyboard (A) | All functionality (select patient, filter, sort, toggle mode, open provenance) is operable by keyboard alone. |
| 2.1.2 | No Keyboard Trap (A) | Keyboard focus can move away from any component without a trap. |
| 2.4.3 | Focus Order (A) | Focus order is logical and preserves meaning (overview -> filters -> list -> detail). |
| 2.4.7 | Focus Visible (AA) | Keyboard focus indicator is always visible. |
| 2.4.11 | Focus Not Obscured (Minimum) (AA, new in 2.2) | A focused element is not entirely hidden by other content (e.g. a sticky header covering the focused list row). |
| 2.5.8 | Target Size (Minimum) (AA, new in 2.2) | Interactive targets are at least 24x24 CSS px (or have adequate spacing). |
| 3.1.1 | Language of Page (A) | The page declares its language (lang attribute). |
| 3.2.4 | Consistent Identification (AA) | Components with the same function are identified consistently (state chips, bars use the same encoding everywhere). |
| 3.3.2 | Labels or Instructions (A) | Labels/instructions are provided for the search and filter controls. |
| 4.1.2 | Name, Role, Value (A) | UI components expose name/role/value to assistive tech (buttons, listbox options, toggle state). |
| 4.1.3 | Status Messages (AA) | Dynamic status changes (filter result count, selected patient) are announced without moving focus (aria-live region). |

**Table 10.** Nielsen heuristic evaluation (10 heuristics).

| # Heuristic | Verdict | Note |
| --- | --- | --- |
| 1. Visibility of system status | MET | aria-live filter count, selected-row highlight + aria-selected, state banner always visible. |
| 2. Match between system and real world | MET | Clinical axis names + units; community mode uses lay language; growth-chart-style centile metaphor. |
| 3. User control and freedom | MET | Filters toggle off, search clears, mode reversible, provenance dialog closes; no destructive actions. |
| 4. Consistency and standards | MET | One state encoding (colour+glyph+word) everywhere; WCAG/ARIA patterns; consistent bar semantics. |
| 5. Error prevention | MET | Read-only viewer, no data entry beyond search; no invalid states reachable; abstain/refuse prevent over-description. |
| 6. Recognition rather than recall | MET | Filters are visible chips; sort is a labelled select; overview shows the population before drill-in. |
| 7. Flexibility and efficiency of use | PARTIAL | Keyboard arrows + search + sort for experts; no saved views / multi-select / bulk export yet. |
| 8. Aesthetic and minimalist design | MET | One patient at a time; progressive disclosure (strata behind details, provenance behind dialog). |
| 9. Help users recognise/ diagnose/ recover from errors | PARTIAL | Empty-filter state shows 0 count but no 'no matches - clear filters' affordance. |
| 10. Help and documentation | PARTIAL | Inline hints + accountability line + provenance; no dedicated help panel / legend for glyphs. |

This work is a conformance and heuristic audit conducted by the developer; a clinical user study with real clinicians and patients remains future work. The three visible states - describe, abstain and refuse - and the hospital and community language registers are shown in Appendix A (Figures A3-A6).

## 4 Discussion

PORTRAIT answers a specific clinical question: where does one individual sit against a reference population across twelve cardiometabolic markers, and how much of that position can be stated with calibrated confidence? The tool establishes a usable operating mode that is narrow and coverage-checked, and defined as much by where it declines to speak as by where it does.

The describability gate checks group-conditional coverage on held-out data before any coordinate is reported; this is a group-conditional empirical check on pre-specified strata, not pointwise conditional coverage for an individual. Per-slice empirical coverage was held inside the design band on 4 of 5 seeds with abstention at 0.101, and a covariance-matched null was refused (anti-cheat pseudo-R2 0.025) rather than passing on surface geometry. The higher-power replication (n = 11668) never under-covered, and where it missed the symmetric-band criterion it erred toward over-covering, the correct failure direction for a coverage check that gates downstream description [12,13].

The deployment argument follows from a second finding. On a genuinely disjoint cohort (NHANES cycle G, n = 2247) a local 50/50 split under-covered all six age-by-sex strata, reaching 0.71, while freezing the reference at n = 9421 placed every stratum back inside the calibrated band and simultaneously lowered abstention from 0.227 to 0.113. The mechanism is calibration depth. A per-cohort split estimates each stratum’s conformal quantile on roughly 180 rows, so small strata under-cover with large finite-sample variance [12,13]; a deep frozen reference estimates the same quantiles once and transfers them. The deployment prescription is therefore to fit the reference once, score each individual against it, and not re-split per cohort, because re-splitting starves per-stratum calibration exactly where individuals are scored one at a time [1]. This is why a frozen background is shipped rather than a per-deployment split.

The interpretation of profile coherence requires the most caution. Coherence is a descriptive coordinate, not a detector. It is only moderately orthogonal to raw extremity (Spearman 0.444), it is not recoverable from centroid distance (R2 0.025), and it correlates empirically with a copula-Mahalanobis distance at Spearman 0.892. It is deliberately not identical to that distance and is not the squared Mahalanobis distance or a chi-square-d statistic; the non-redundancy is exactly what lets it add per-feature information. Against a clean null it concentrates and carries no detection advantage over Mahalanobis. What it adds is attribution: per-feature person-fit residuals surface jointly implausible combinations that a marginal read passes. The worked exemplar illustrates this. A fasting glucose at the 0th centile (66 mg/dL) paired with a mid-range HbA1c is physiologically discordant, flagged by a residual of r = −3.84 given the rest, while each analyte read alone raises no alarm. The informative part is the attribution, not the scalar magnitude. The exploratory standout analysis fixes the boundary: apparent clinical associations of coherence are repackaged raw extremity. Every clinical marker that looked associated at M1 collapsed once per-feature surprise was adjusted at M2, and the robust carrier is the ordinary surprise count, which shows a monotone mortality dose-response (HR 1.292 per surprising feature). Coherence is therefore not an independent clinical axis. One residual finding is a fairness signal: coherence associates with family poverty-income ratio (beta −0.472, BH q = 0.079), reported to monitor in deployment, not to act on [22,23].

The mortality increment deserves the same discipline. The mortality-endpoint analysis found a real, permutation-verified gain of deltaC = 0.0047 over the marginal features plus age and sex, against a baseline C of 0.8289, holding under frailty adjustment. This is positive but clinically negligible, a few thousandths of concordance at the analytic-sample level, and the standout reconciliation shows it is largely repackaged extremity: the continuous coherence hazard ratio attenuates from 1.388 to 1.231 once surprise is added, and the binary flag does not survive at all (HR 0.93, p = 0.80). It is stated as a small, mostly repackaged increment, not an independent prognostic contribution [15,16].

The interface is a first-class contribution, not packaging. An evidence-grounded, abstention-first Passport matters because the dominant deployment risk is not a wrong number but an over-trusted one. Clinicians and lay users over-rely on usually-accurate tools and will reverse correct decisions when the tool errs [7]; the mitigations with the strongest support are explicit accountability cues and confidence attached to output [7], both of which are built in through the standing responsibility line and the three visible states. The reject option that abstains when likely to err is the ABSTAIN state, which passes the undescribable case back to the clinician by design [2]. Uncertainty presentation follows the graded evidence: natural-frequency glosses and horizontal reference-range bars with goal anchors, because frequency framing improves probabilistic inference and harm-anchored bars reduce overestimated urgency of out-of-range values [6,9,28,29]. The WCAG 2.2 audit, with state carried by label and glyph rather than colour alone, aria-labelled bars, and scoped-header tables, is what makes the descriptive claim survive contact with real users. On regulatory readiness the posture is honest: PORTRAIT is framed against DTAC classification, DCB0129/DCB0160 clinical risk management, and IEC 62366-1 usability engineering, and the byte-deterministic render, tying each Passport to its frozen reference and code hash, supports that framing. Several regulatory items remain unmet and require formal processes: a Clinical Safety Officer, a completed hazard log and safety case, and full usability validation.

The contribution is the combination: a coverage-checking describability gate, a principled refusal that abstains rather than guesses, a calibrated joint coordinate with per-feature attribution, and an accessible interface, evaluated on a frozen population reference. Each piece exists somewhere in the literature; assembling them into a single honest, calibration-checked description is what is new. The coherence scalar is, by construction, closely related to a known distance, and that is stated plainly; its value is the attribution it exposes, not novelty as an estimator.

This closes the arc. Because PORTRAIT is calibrated and refuses when it cannot describe, it can sit behind a single accessible interface intended for use in hospital, general practice, or a high-street setting. The impact is concrete. A clinician, or a high-street user, sees a calibrated position, the features that drive it, and an explicit refusal when the tool cannot support a statement, instead of an over-confident single number. PORTRAIT is a descriptive tool: it reports where an individual sits and where their profile conflicts, within the preconditions where the gate’s coverage check is met, and it hands back a case that failed the check explicitly rather than a confident-looking artefact.

### 4.1 Clinical utility: a staged evaluation programme

Calibration and abstention are necessary but not sufficient for clinical utility: they establish that when PORTRAIT speaks it is statistically honest in a well-defined sense, not that speaking changes a decision for the better. For a tool that describes and abstains rather than predicts, utility is a question of process and decision quality, not of discrimination. The evaluable claims are specific: that a reader is more likely to recognise a clinically relevant discordance between results; that fewer automation-biased errors are made than with an always-answer tool; that a lay reader’s understanding improves without inappropriate reassurance or alarm; and that follow-up is neither missed for a discordant profile nor triggered unnecessarily for a benign one. The present NHANES results establish a well-behaved descriptive spine; the evidence for those utility claims requires a staged programme, summarised here as future work.

The near-term, most defensible stages are retrospective adjudication and controlled human-factors experiments. Expert panels, blinded to the PORTRAIT screen, would rate stratified sets of profiles - conventionally normal, high-coherence, high conditional-surprise, and abstained - for whether they warrant investigation, reassurance, or suspicion of artefact, and these ratings would be compared against the tool’s states to test alignment with expert judgement; convergence with established risk stratification (for example QRISK3 [5]), adjusted for age, sex and marginal centiles, would be examined as convergent evidence rather than new prediction.

Randomised clinician vignette experiments would then present the same profiles under three display conditions - a standard laboratory report with reference ranges and high/low flags, the PORTRAIT descriptive screen, and a naive always-answer score with no abstention - against a ground-truth good-decision set defined independently of PORTRAIT’s signals. The outcomes are detection of clinically relevant discordance, the rate of unsafe decisions, direct measures of automation bias and of response to the abstention state [2,7], and time-on-task and workload.

Longer-term stages move into workflow. A shadow-mode deployment would measure coverage and abstention in a real clinical population rather than NHANES, quantify how often the three states diverge from conventional interpretation, and audit the cases of strong disagreement for latent issues such as mis-coded units or missed follow-ups. A single-centre before-and-after introduction would compare process measures - follow-up tests ordered for discordant profiles, time to consultant review, documentation of conflict, artefact corrections - treated as hypothesis-generating given confounding by temporal trend and case mix. Patient-facing utility requires its own evidence: comprehension experiments comparing the community view against standard result letters, on understanding, appropriate reassurance, and the distinction between the reference population, healthy, and the individual, alongside qualitative work on emotional impact and trust [8,10,26,28,29]. Only after these would a pragmatic cluster-randomised or stepped-wedge trial be warranted, reported to the DECIDE-AI and CONSORT-AI standards for early-stage and trial-stage clinical AI, and with process endpoints - detection of important discordance leading to appropriate follow-up, reduction in inappropriate reassurance - rather than mortality or long-term outcomes, for which the tool is deliberately not designed.

This programme belongs to a separate clinical-utility study; the present paper establishes the calibrated, abstention-aware descriptive method on which it builds. Should any predictive or prognostic claim be added in future, the appropriate reporting frameworks would be TRIPOD+AI and, for diagnostic-index use, STARD-AI; the present descriptive-methods manuscript is deliberately not forced into a prediction-model framework.

## 5 Limitations

### Several constraints bound what this work supports

First, all NHANES analyses are unweighted. The analytic sample is treated as a fixed finite population; centiles and coverage are within-sample descriptive quantities. Population-representative inference requires the survey weights, which were not applied, so no claim here generalises to the US population. Relatedly, two markers use the MEC weight as an approximation for the fasting subsample.

Second, distribution-free conditional coverage is impossible [11]. The gate therefore provides group-conditional coverage over fixed, pre-specified slices only, and this guarantee degrades as a stratum shrinks, the frozen-versus-split lesson stated above. Coverage statements are marginal within each declared slice, not per-individual, and thin strata carry correspondingly weak assurance.

Third, transfer has preconditions and is not universal. Two negative controls show this directly: the ocular transfer did not hold coverage at thin-n (myopia, n_train 309), and orthogonality failed under collinear features (biometry, 0.27 and 0.67, against 0.44 in the primary setting).

PORTRAIT requires adequate calibration depth and approximately orthogonal coordinates; where these fail, it should not be deployed.

Fourth, the mortality signal is small. The C-index increment (deltaC 0.0047 [23]) is analytic-sample-level and largely repackaged extremity. It is a descriptive association, not a clinical prediction or risk claim, and should not be read as prognostic performance.

Fifth, the standout analysis is exploratory. It rests on a single cohort, is unweighted, and the flagged group is small (n = 115). The coherence-to-poverty-income fairness signal is a hypothesis-generating observation and needs weighted confirmation in independent data before any interpretive or operational use.

Sixth, the interface evaluation is a single-site heuristic walkthrough plus a conformance audit conducted by the developer. It is not a clinical user study with real clinicians or patients, and it does not establish resistance to automation bias or safe risk communication in practice [7,29].

The remaining regulatory and human-factors engineering requirements are unmet pending formal summative processes; the present audit is a precondition, not a substitute.

Seventh, coherence correlates with the copula-Mahalanobis quantity at Spearman 0.892 but is deliberately not identical to it [16]. It cannot, and is not intended to, outperform that quantity as a detector; any apparent clinical signal is extremity re-expressed. Its contribution is descriptive attribution, locating and naming where a profile sits, not detection.

Eighth, the reference is US NHANES. Deployment to other populations, including UK settings, is a reference-portability question. The reference is swappable, but it must be re-frozen and re-validated on the target population before use. This is scope, not a defect.

Ninth, the portrait is built on twelve routine features in adults only. Other feature sets, alternative measurement panels, and paediatric reference ranges are untested, and the descriptive coordinates should not be assumed to behave equivalently outside this scope.

Taken together, these limitations mark PORTRAIT as a descriptive coordinate system with known failure modes, not a diagnostic, predictive, or population-inference tool. The negative controls and the frozen-versus-split analysis make its boundaries visible.

## Data Availability

GitHub

https://github.com/doehring-gh/PORTRAIT

## Declarations

### Ethics

This study used de-identified public data (NHANES and its CDC-linked mortality files) and a restricted dataset (MIMIC-IV) accessed under the PhysioNet Data Use Agreement for a local counter-test only. No identifiable data were used and no new data were collected from human participants; institutional ethical review was therefore not required.

### Data availability

NHANES cycles D-H are public (CDC/NCHS), with download URLs in the repository; the myopia dataset is public; MIMIC-IV was used only for a local counter-test and is not redistributed, under the PhysioNet DUA; CDC NHANES-linked mortality is public with usage terms, with a fetch recipe and expected hashes in the repository and the file not redistributed.

Code and derived results are in the public repository at https://github.com/doehring-gh/PORTRAIT [25], with a DOI on posting.

### Use of AI tools

The analyses, code, and figures were produced with Claude (Anthropic) operating as an autonomous research engineer within the Claude Science platform, under the author’s direction.

The author designed the study, made all aim-level and clinical-framing decisions, reviewed each stage, and takes full responsibility for the content. Every reported number is traceable to a recorded computational environment and content hash. Some AI-designed checks returned negative results, which are reported alongside the positive ones. No patient-identifiable data were provided to any model; restricted data (MIMIC-IV) were never transmitted off the local machine. A full account of how the tool was used is in the repository (CLAUDE_USE_AUDIT.md).

### Author contributions

D.O. conceived the study, directed the analysis, made all aim-level and clinical decisions, approved each phase gate, and wrote and approved the manuscript.

## 6 Appendix A. The PORTRAIT interface

This appendix shows the PORTRAIT application rendered from the frozen held-out cohort (n = 2248). Each figure is a screenshot of the actual single-file browser application; no statistics are computed in the page. The interface is described in Methods (The clinical interface) and evaluated in Results (Interface evaluation).

**Figure A1.**
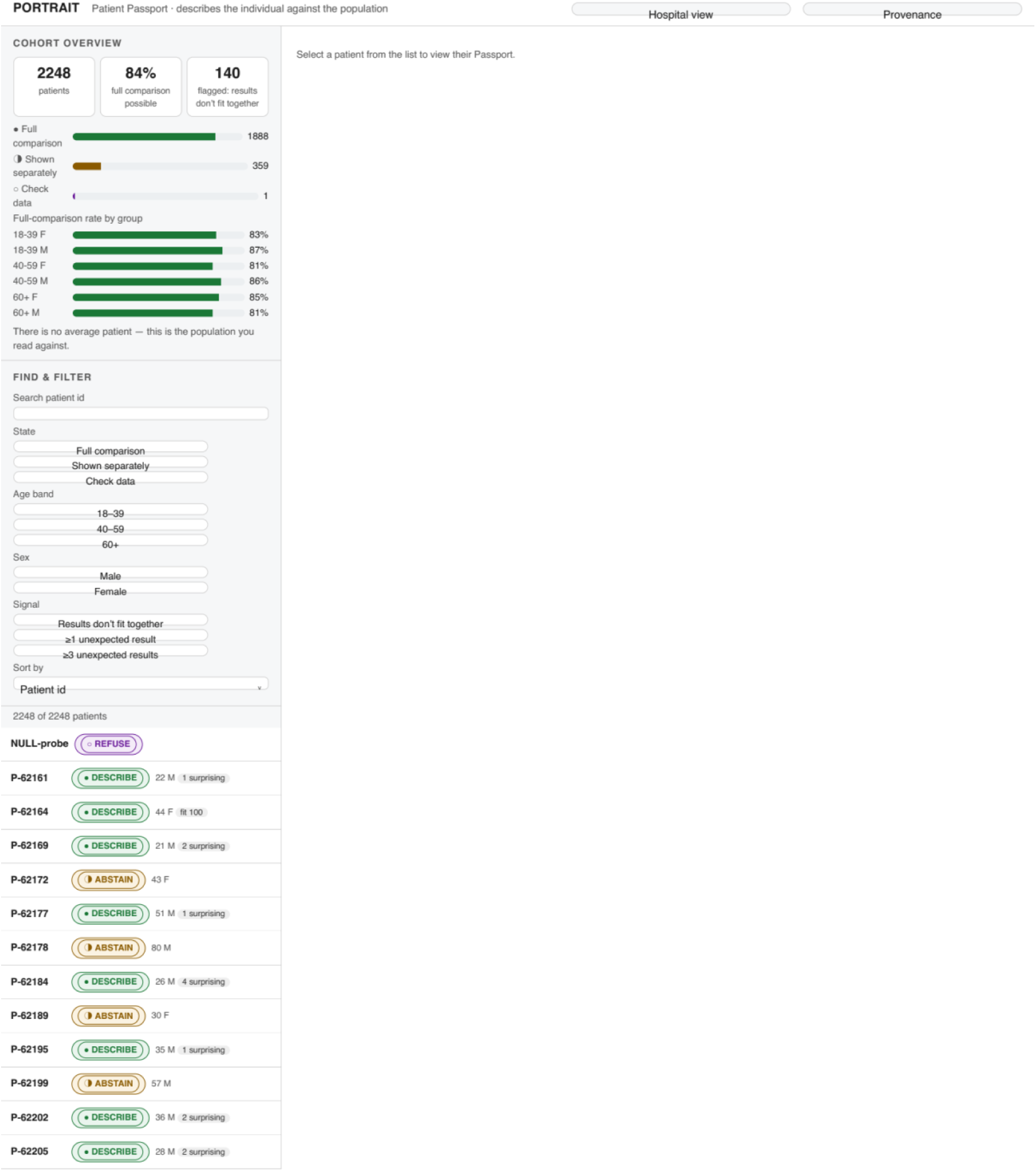
Cohort overview and the patient list. The opening view. The left rail shows the cohort overview - total patients, the proportion for whom a full comparison is possible, and the count flagged as results-do-not-fit-together - followed by per-group full-comparison rates and the find-and-filter controls. The right pane is empty until a patient is selected. Every patient in the held-out cohort (n = 2248) is listed with a state chip; the single REFUSE record (NULL-probe) sorts to the top. Typing in the search box, pressing a state or age-band or sex chip, or choosing a signal filter narrows the list in place; the count above the list updates live.

**Figure A2.**
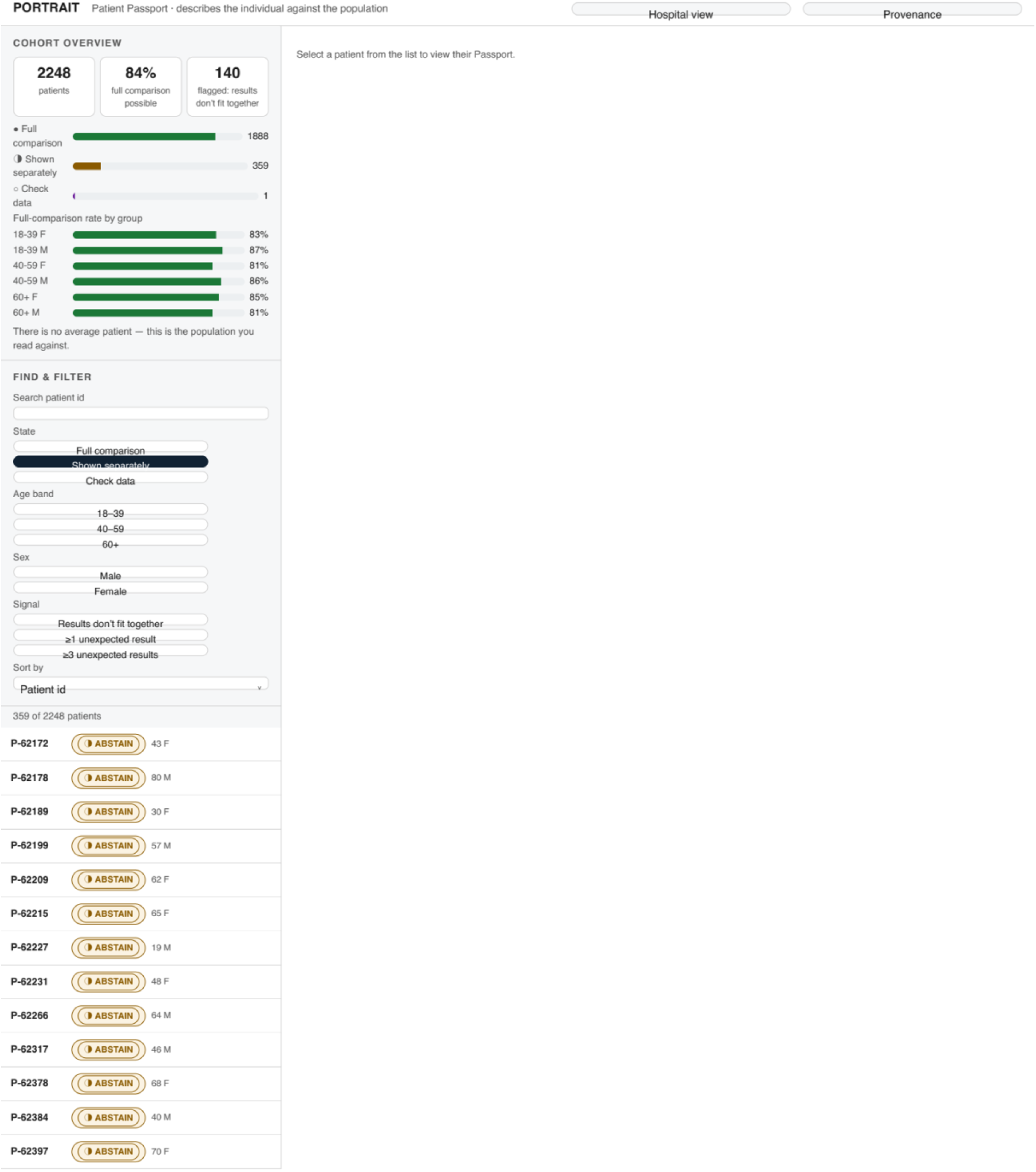
Filtering to the abstained records. The result of pressing the Shown separately state chip. The list is reduced to the 359 abstained records and the live count reads 359 of 2248 patients. Filters combine: adding an age band or a signal chip intersects the criteria. This is how a clinician isolates, for example, all patients for whom the tool declined a joint description within a given age group.

**Figure A3.**
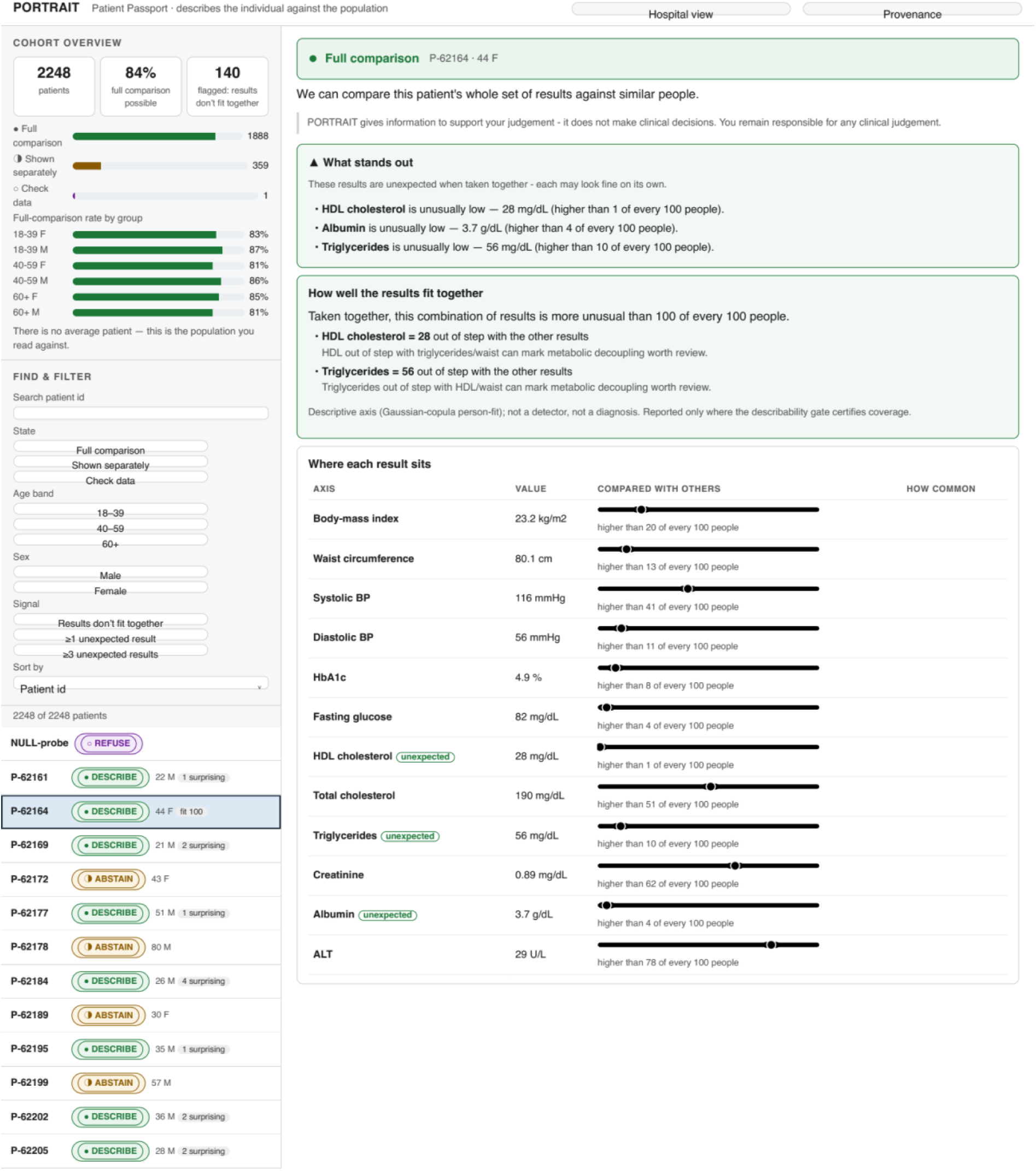
A described patient’s Passport (hospital view). Clicking a patient row opens their Passport in the right pane. Patient P-62164 (44 F) is in the Full comparison (DESCRIBE) state. The Passport shows, in order: the state banner and the accountability line; What stands out, listing the features that are unexpected given the rest of the profile (here HDL cholesterol, albumin and triglycerides, each unusually low); How well the results fit together, the coherence coordinate with its per-feature drivers; and Where each result sits, a table giving every analyte’s value, its position against the population as a labelled bar, and a natural-frequency gloss. This is the worked example discussed in the main text: each value looks individually plausible, yet the combination is flagged.

**Figure A4.**
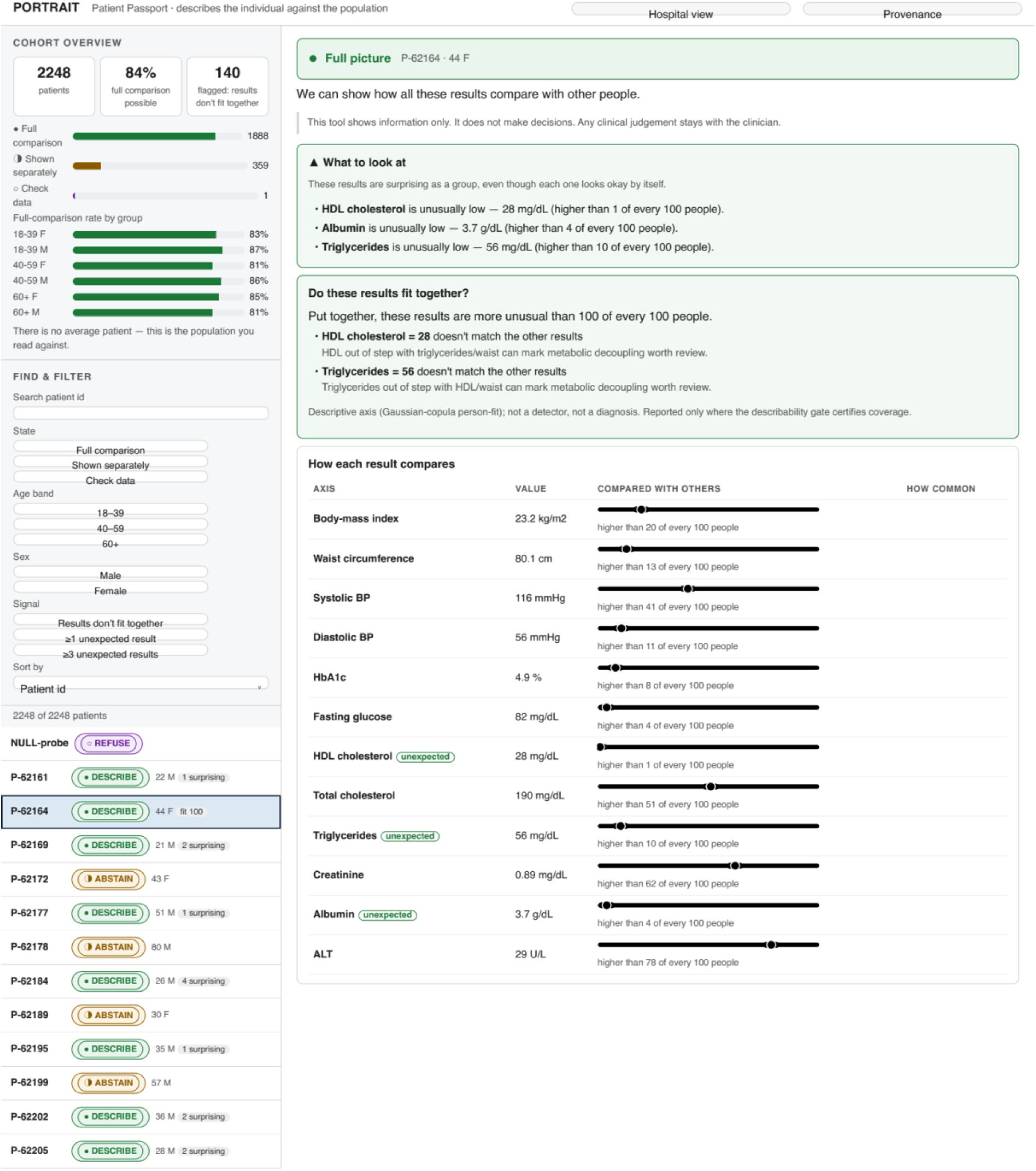
The same Passport in community (high-street) view. Pressing the Hospital view / Community view toggle in the header re-renders the same patient with plainer language while the underlying numbers are unchanged. Full comparison becomes Full picture; the lead sentence and section headers switch to lay phrasing. The toggle changes wording only - no statistic is recomputed - demonstrating the single-render, dual-register design intended for both hospital and high-street settings.

**Figure A5.**
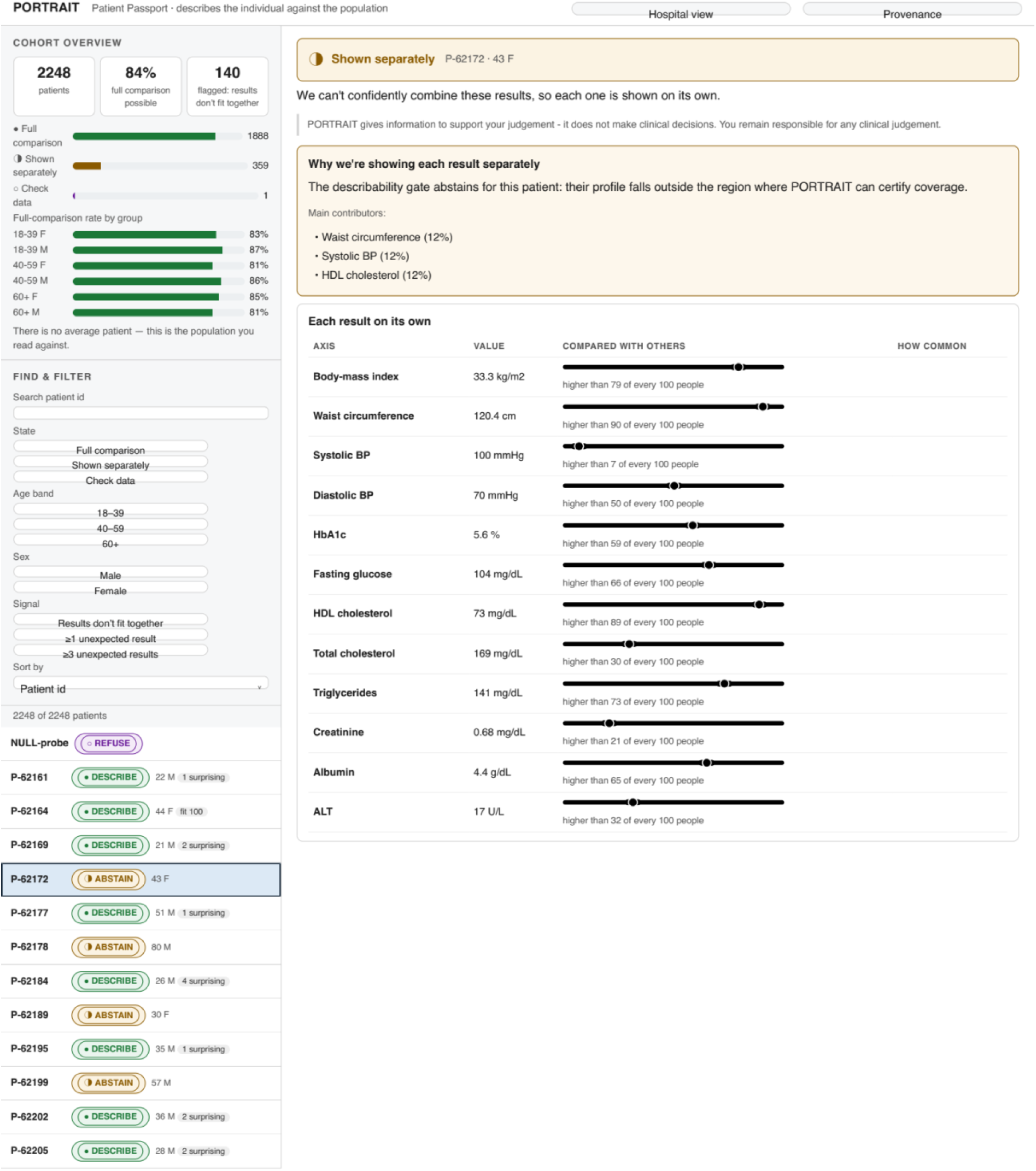
An abstained patient’s Passport. Patient P-62172 is in the Shown separately (ABSTAIN) state: the frozen reference cannot meet the coverage check for a calibrated joint description of this record, so the tool declines to combine the results and instead presents each analyte on its own, with an explanation of why. This is the visible refusal that distinguishes PORTRAIT from displays that always produce an output.

**Figure A6.**
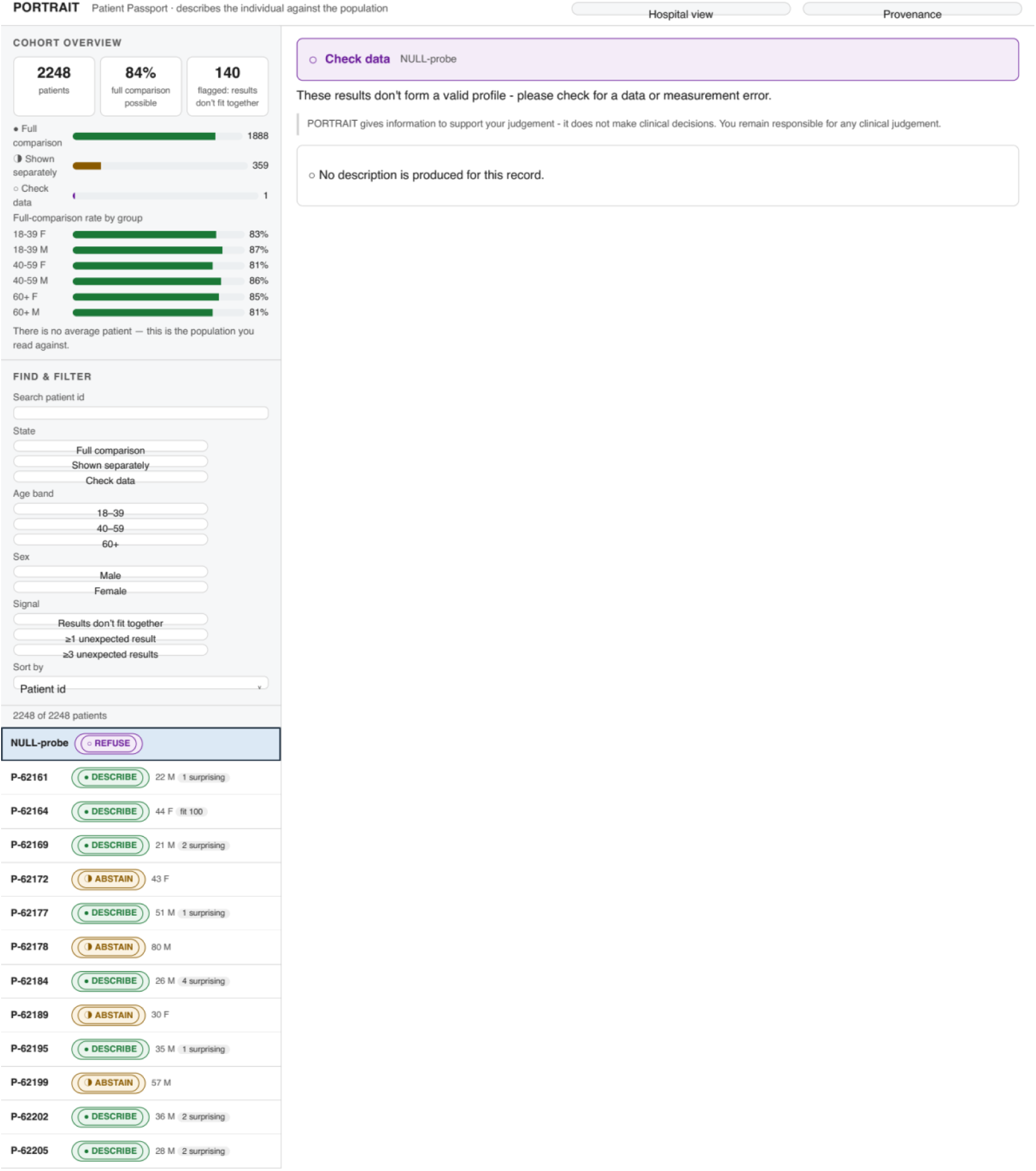
A refused record. The NULL-probe record is in the Check data (REFUSE) state. The profile does not form a valid input, so no description is produced and the interface asks the reader to check for a data or measurement error. REFUSE is a distinct, deliberate state - not a silent failure or a default value.

